# Cholesterol-lowering Decreased mTOR Complex 2 Signaling and Enhanced Antitumor Immunity

**DOI:** 10.1101/2021.01.29.21250381

**Authors:** Yanping Wang, Sungyong You, Shengchen Su, Austin Yeon, Eric M. Lo, Sungjin Kim, James L. Mohler, Michael R. Freeman, Hyung L. Kim

## Abstract

Cholesterol-lowering interventions are employed widely and safely to reduce risk of cardiovascular disease. Cholesterol may have complex and opposing effects on immunity. We lowered serum cholesterol to clinically relevant levels in mice and evaluated the final adaptive immune response. Mice treated with oral ezetimibe exhibited enhanced antitumor immunity against syngeneic cancers in a CD8+ lymphocyte-dependent manner, produced immunity that was transferrable through lymphocytes, and enhanced central CD8+ T cell memory. In both mice and patients undergoing prostatectomy, lowering serum cholesterol inhibited mTORC2 signaling in lymphocytes and increased infiltration of CD8+ lymphocytes into prostate tumors. Lymphocyte-specific mTORC2 knockout mice demonstrated enhanced CD8+ lymphocyte function and antitumor capacity. In a prospective clinical trial, cholesterol-lowering intervention prior to prostatectomy decreased the proliferation of normal prostate and low-grade adenocarcinomas. Here, we show that lowering serum cholesterol may be an effective strategy to decrease signaling through mTORC2 and enhance antitumor CD8+ T cell memory.

## INTRODUCTION

Prostate cancer is the most commonly diagnosed cancer and the second leading cause of cancer death among men in North America.^1^ In the U.S., there were 174,650 newly diagnosed cases and 31,620 deaths from prostate cancers annually. Over the past 10 years a large body of evidence has emerged from pre-clinical ^2, 3, 4, 5^ and observational studies that cholesterol-lowering (CL) HMG-CoA-reductase inhibitors (i.e., statins) exert a protective effect against prostate cancer.^6^ A meta-analysis of 15 cohort and 12 case-control studies concluded that statin use was associated with a reduction in total incidence of prostate cancer and a larger reduction in advanced prostate cancer.^7^ The possibility of repurposing a widely used cardiac drug with a well-established safety profile to treat prostate cancer, and possibly other cancers, is highly attractive.

The potential molecular mechanisms underlying the oncologic benefits of CL have been explored. Cholesterol-lowering drugs have been proposed to directly alter tumor growth^3, 5, 8^. Cholesterol-manipulation may modulate T cell immunity, which is largely responsible for immune-mediated cancer control.^9, 10, 11^ However, previous reports that examined the immune effects of CL at the subcellular level have reached conflicting conclusions. Some reports suggest that membrane cholesterol promotes T cell function. Yang et al. reported that ACAT1-deficient CD8+ T cells, which have increased membrane cholesterol, showed enhanced effector function and improved control of melanoma growth and metastasis in mice.^12^ Consistent with this model, Wang et al. reported that cholesterol sulfate, a naturally occurring analog of cholesterol, inhibits T cells by disrupting cholesterol needed for activation of T-cell receptors.^13^ However, a report by Swamy et al. suggests that membrane cholesterol inhibits T-cells because only T-cell receptors that spontaneously detach from cholesterol could activate and signal.^14^

These studies relied on *in vitro* methods and genetic strategies that manipulated cholesterol levels in specific immune cells. However, cholesterol is an essential structural component of all cell membranes, and lowering serum cholesterol may have varied, and even opposing, antitumor effects that depend on immune context and cells studied. Sag et al. reported that when the cholesterol transporter ABCG1, which is responsible for efflux of cholesterol from cells, was knocked out, antitumor immunity was enhanced because there was a bias toward formation of tumor-fighting M1 macrophages.^15^ In contrast, Ma et al. reported that lowering cellular cholesterol content favored polarization of CD8+ T cells to IL9-secreting cells rather than classic Tc1 cells, and this led to a stronger antitumor response against mouse melanoma.^16^ Therefore, based on the subset of the immune system being assessed, cholesterol may appear to have contrasting effects on antitumor immunity.

Therefore, to translate CL as a strategy for immune modulation in patients, we used preclinical models where it was possible to selectively lower serum cholesterol and assess the final antitumor immunity resulting from the varied effects of CL on all components of the immune system. Prior attempts to do this in mouse models have relied on statins. Statins are the most commonly prescribed drug for CL. In patients, statins’ primary function is to inhibit cholesterol synthesis in the liver. However, statins have a modest effect on mouse serum cholesterol and may increase cholesterol synthesis in the mouse liver.^17, 18^ Furthermore, statins have effects in mice that are unrelated to changes in serum cholesterol.^17^ For example, statins decrease isoprenoids and subsequent prenylation, which are effects not mediated through cholesterol.^19^ Therefore, we used oral ezetimibe to inhibit the intestinal cholesterol absorption and reduce serum cholesterol in mice by levels analogous to statin-treated patients.^3, 20^ An import feature of ezetimibe is that it remains in the gut and is not systemically absorbed; therefore, it does not have any direct effects on the tumor or immunity.^21^

In this work we characterize the adaptive immunity in a preclinical model where serum CL is verified. We show that CL in mice decreases the growth of melanoma, prostate cancer and kidney cancer cells. To establish the role of the immune system, we show that inhibition of tumor growth is CD8+ lymphocyte-dependent. Additional evidence of immune mediated antitumor effects is provided by adoptively transferring lymphocytes from mice subjected to CL. These lymphocytes are able to prevent growth of metastatic tumor deposits in mice that did not receive any other treatment. By monitoring Pmel-1 cells adoptively transferred to B6 mice, we show that CL enhanced central CD8+ memory cells, providing a mechanism of action. We show in mice and humans that lowering serum cholesterol inhibits the AKT pathway in lymphocytes, increases infiltration of CD8+ lymphocytes in the tumor, and decreases proliferation of tumor cells. We show that these effects are mediated through the mTORC2 pathway in lymphocytes. CL interventions are widely and safely used to reduce the risk of cardiovascular disease in patients and the potential to modulate immunity by simply lowering serum cholesterol has large public health implications.

## RESULTS

### Cholesterol-lowering enhances antitumor immunity

We’ve previously reported that CL interventions reduce the growth of murine prostate cancers by inhibiting mTOR signaling.^5^ We and others have reported that mTOR inhibition can promote the formation of CD8+ lymphocyte memory formation, particularly central memory formation, and this can lead to enhanced antitumor immunity.^22, 23, 24^ Therefore, we hypothesized that CL may lead to antitumor immunity mediated by CD8+ lymphocytes. To test the possibility that lowering serum cholesterol may enhance antitumor immunity, we treated mice with ezetimibe, which lowers cholesterol by binding to the intestinal cholesterol transporter NPC1L1, thus preventing cholesterol absorption.^21^ Ezetimibe is not absorbed from the gut into the blood stream, and therefore, cannot have a direct effect on the tumor. In three different syngeneic mouse tumor models (melanoma, prostate cancer, and renal cell carcinoma), ezetimibe decreased tumor growth **(Fig. 1A-D)**. This was at least in part immune-mediated, since depletion of CD8+ lymphocytes negated the majority of the antitumor effects of ezetimibe.

**Figure 1.**
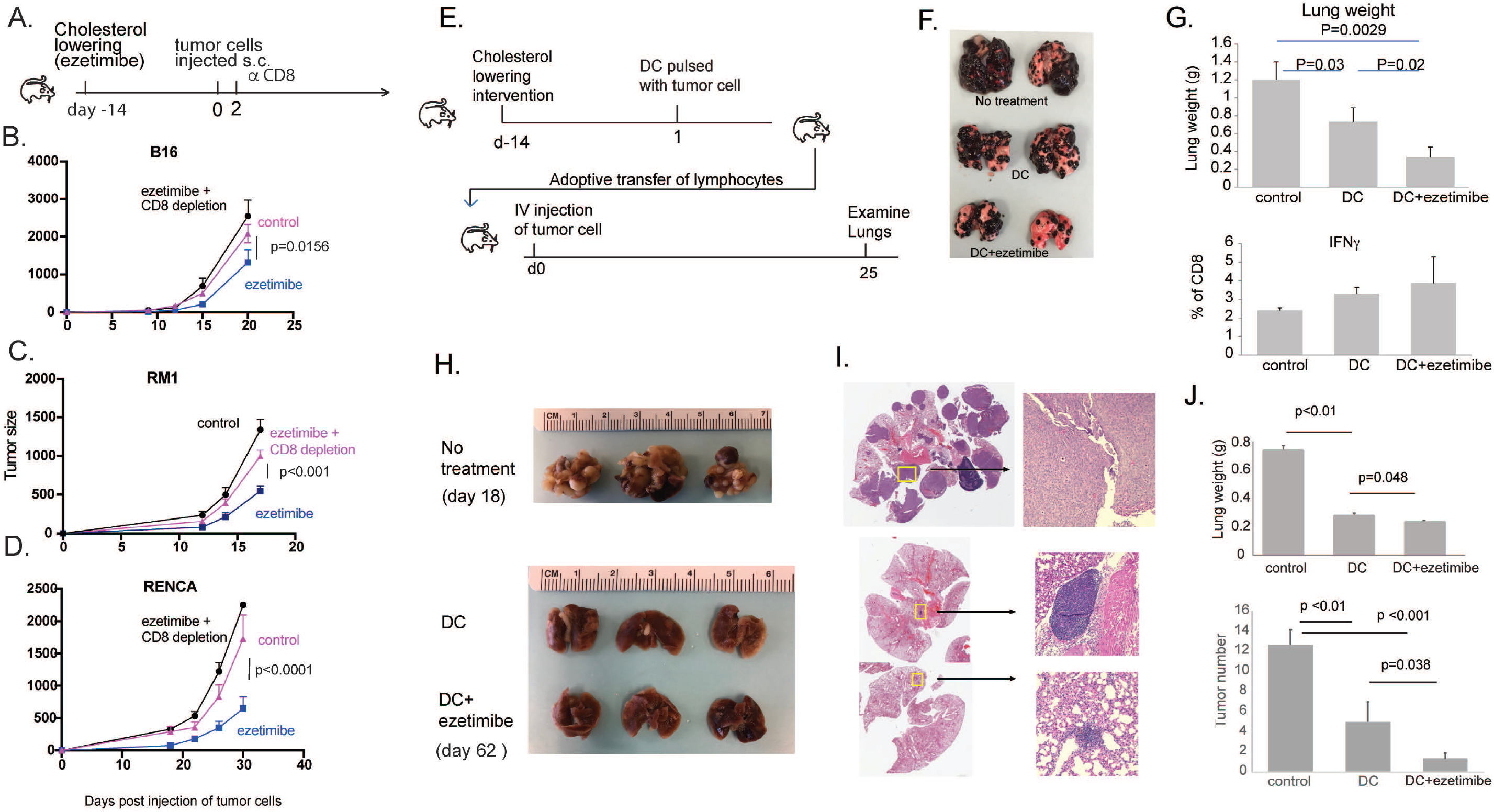
Ezetimibe enhances lymphocyte function to inhibit tumor growth and metastasis in mice. **A.** Mice were treated with ezetimibe for 14 days prior to subcutaneous injection of syngeneic tumor cell lines. **B**. B16 melanoma cells were injected into B6 mice (8-9 mice/group) **C**. RM1 prostate adenocarcinoma cells were injected into B6 mice (9-10 mice/group). **D**. RENCA renal cell carcinoma cells were injected into Balb/c mice (5-6 mice/group). In each experiment, one group received ezetimibe and depleting αCD8 antibody two days after s.c. tumor injection into the flank. P-values were obtained by using the repeated measures ANOVA (A-D). **E**. To determine if immunity is lymphocyte-dependent, mice were treated with ezetimibe for 14 days to lower cholesterol and then vaccinated with DCs pulsed with B16 tumor lysate. Lymphocytes were harvested from these mice and cultured *in vitro* with tumor lysate pulsed DC and IL-2 for 3 days, and adoptively transferred into recipient mice, which were challenged with i.v. B16 cells. Lung tumors were examined 25 days later. **F**. Representative lungs are shown from recipient mice, labeled with the donor mouse treatment. **G**. Lung weights (top) and CD8+ T cell IFN-*γ* response to *ex vivo* restimulation (bottom) are shown. Graphs are labeled with the treatment provided to the donor mice. **H**. In a similar experiment, B6 mice were treated with DCs pulsed with RM1 tumor lysate. Lymphocytes were transferred into recipient mice that were challenged with i.v. RM1 cells. **I**. Representative H&E sections of lungs from each treatment group are shown. **J**. Lung weights (top) and tumor number in the lungs (bottom) are shown. Each group contained 3 mice. Each experiment was conducted at least twice to confirm results (E-J).

To provide definitive evidence that the antitumor effect associated with CL was mediated by adaptive immunity, we performed a lymphocyte transfer study **(Fig. 1E)**. Mice treated with ezetimibe to lower serum cholesterol were administered a cancer vaccine consisting of dendritic cells (DC) pulsed with B16 tumor lysate. Lymphocytes were harvested from these mice 7 days later and adoptively transferred into na^ï^ve mice (i.e. mice were never subjected to CL), which then were challenged intravenously with B16 tumor cells. The tumor cells were administered directly into the tail vein; this model represents a very aggressive cancer scenario where only the most potent treatments are expected to show benefit. Compared to control mice that received lymphocytes from untreated mice, mice that received lymphocytes from vaccine-treated donor mice had fewer lung tumor deposits, and those that received lymphocytes from mice treated with vaccine plus ezetimibe had the fewest lung deposits **(Fig. 1F)**. Consistent with this observation, mice in the latter group with smallest tumor burden had the lowest lung weight. In addition, the CD8+ lymphocytes retrieved from these mice tended to have a greater interferon-*γ* response upon restimulation with DC pulsed with B16 tumor lysate **(Fig. 1G)**.

These findings were reproduced using RM1 prostate cancer cells. Mice with lymphocytes transferred from mice treated with vaccine plus ezetimibe had the fewest tumor deposits in the lung **(Fig. 1H,I)** and the lowest lung weight **(Fig. 1J)**. In another version of this experiment, stimulated lymphocytes were transferred to naive mice before they were challenged with subcutaneous RM1 tumor. Mice that received lymphocytes from donor mice subjected to CL had the slowest tumor growth **(S. Fig. 1)**. These results clearly showed that lowering serum cholesterol stimulated a lymphocyte-mediated antitumor immune response that was independent of any direct effects on the tumor cells. Use of ezetimibe instead of a statin, which can have cholesterol-independent effects, allowed us to attribute all immune effects to CL. *In vivo* tumor growth was reduced by the sum of all immune effects resulting from serum CL.

### CL decreases mTORC2 signaling in lymphocytes and enhanced CD8 memory

We confirmed that ezetimibe decreased serum cholesterol in mice **(Fig. 2A)**. The resulting lymphocytes had decreased mTOR signaling in splenocytes (**Fig. 2B)**, including CD8+ lymphocytes **(S. Fig. 2)**. The largest decrease in phosphorylation was seen for AKT, at Ser473, which is associated with mTORC2 signaling. This effect was also observed in patients; pAKT (Ser473) was decreased when comparing lymphocytes before and after 2-6 weeks of CL treatment **(Fig. 2C,D)**. The observation that the mTORC2 pathway may be mediating immune stimulation was surprising because our previous work using first generation mTOR inhibitors showed that antitumor immunity was associated with inhibition of the mTORC1 pathway^25^, and no prior work has linked CL with a decrease in TORC2 signaling.

**Figure 2.**
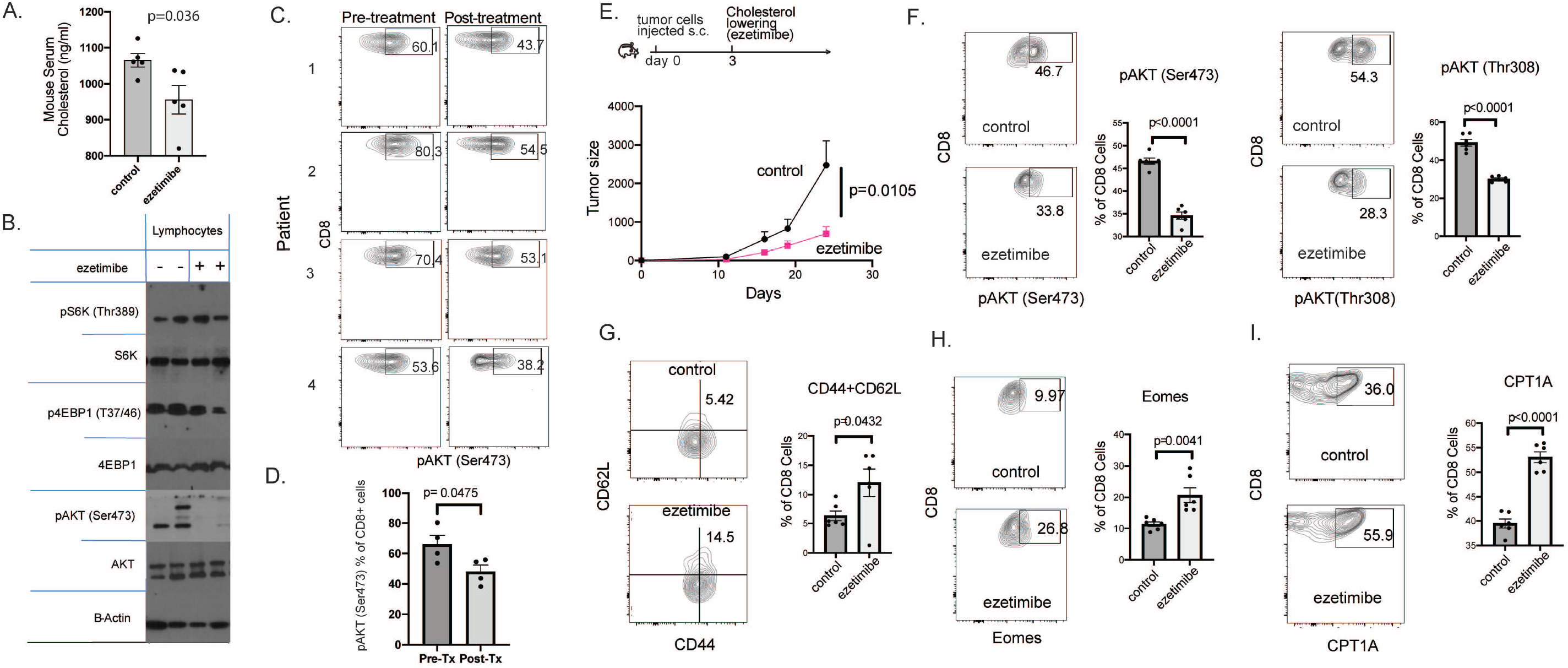
Cholesterol-lowering decreases mTOR signaling in lymphocytes and increased markers for central CD8+ T cell memory. **A**. Serum cholesterol levels were measured in mice treated with ezetimibe (n=6). Splenocytes from ezetimibe-treated mice showed decreased AKT/mTOR signaling, with pronounced decrease in phosphorylation of AKT at Ser473. Representative blots from duplicate experiments are shown. **C**. pAKT (Ser473) in CD8+ lymphocytes from patients before and after cholesterol lowering (CL) with statin+ezetimibe was measured by flow cytometry (n=4). **D**. Human pAKT (Ser473) in CD8+ lymphocytes in these patients was significantly reduced by CL. **E**. RM1 prostate adenocarcinoma cells were injected into B6 mice (8-9 mice/group) and ezetimibe treatments were started 3 days later. Tumor growth was monitored and p-value was obtained by using the repeated measures ANOVA. **F-I**. Lymphocytes were collected from mice treated with ezetimibe or saline for 21 days and analyzed for the indicated markers by flow cytometry. **G**. Splenocytes were first gated for CD8 prior to flow cytometry analysis for the indicated markers.

The experiments in Figure 1 clearly demonstrated that antitumor immunity resulting from CL was lymphocyte dependant. In a slight variation of the experiment in **Figure 1A**, CL was initiated after injection of tumor cells to simulate a clinical scenario where cancer is treated with CL after diagnosis. The ezetimibe-treated mice had decreased tumor growth when compared to control mice **(Fig. 2E)**. In ezetimibe-treated mice, the CD8+ lymphocytes had decreased mTOR2 signaling as measured by decrease in pAKT (Ser473) and pAKT (Thr308) **(Fig. 2F)**. Ezetimibe-treatment was associated with an increase in central memory markers. CD8+ lymphocytes that were CD44+CD62L+ increased **(Fig. 2G)** as well as CD8+ lymphocytes expressing Eomes **(Fig. 2H)**. The mTOR pathway and central CD8+ memory cells have been linked to fatty acid oxidation where the rate-limiting step is mediated by CPT1A.^26^ CL with ezetimibe was associated with increased CPT1A expression in CD8+ lymphocytes **(Fig. 2I)**.

Memory CD8+ T cells mediate long-term antitumor immunity. Decreased mTOR/mTORC1 signaling can trigger the formation of memory CD8+ T cells.^24, 27, 28^ We considered the possibility that CL, with its inhibition of mTORC2, could enhance the formation of memory CD8+ T cells. Thy1.1+ Pmel-1 lymphocytes were adoptively transferred into congenic Thy1.2+ B6 mice **(Fig. 3A)**. Pmel-1 lymphocytes recognize gp100 and can be tracked using Thy1.1. Ezetimibe was administered to lower serum cholesterol, and the mice were injected with bone marrow-derived DCs pulsed with gp100 peptide **(S. Fig. 3)**. In the mice treated with ezetimibe, the percent of donor-derived Pmel-1 lymphocytes on day 10 increased significantly, which indicated an increase in gp100-specific cells **(Fig. 3B, S. Fig. 4)**. Central memory cells have a greater recall response and are able to provide greater protection than peripheral memory cells. Both Thy1.1+ and Thy1.1-CD8+ cells were more likely to have a central memory phenotype (CD62L+CD44+; Eomes+) **(Fig. 3B, S. Fig. 5)**. Consistent with this observation, the Thy1.1+ cells had higher intensity of CPT1A expression **(Fig. 3C)**.

**Figure 3.**
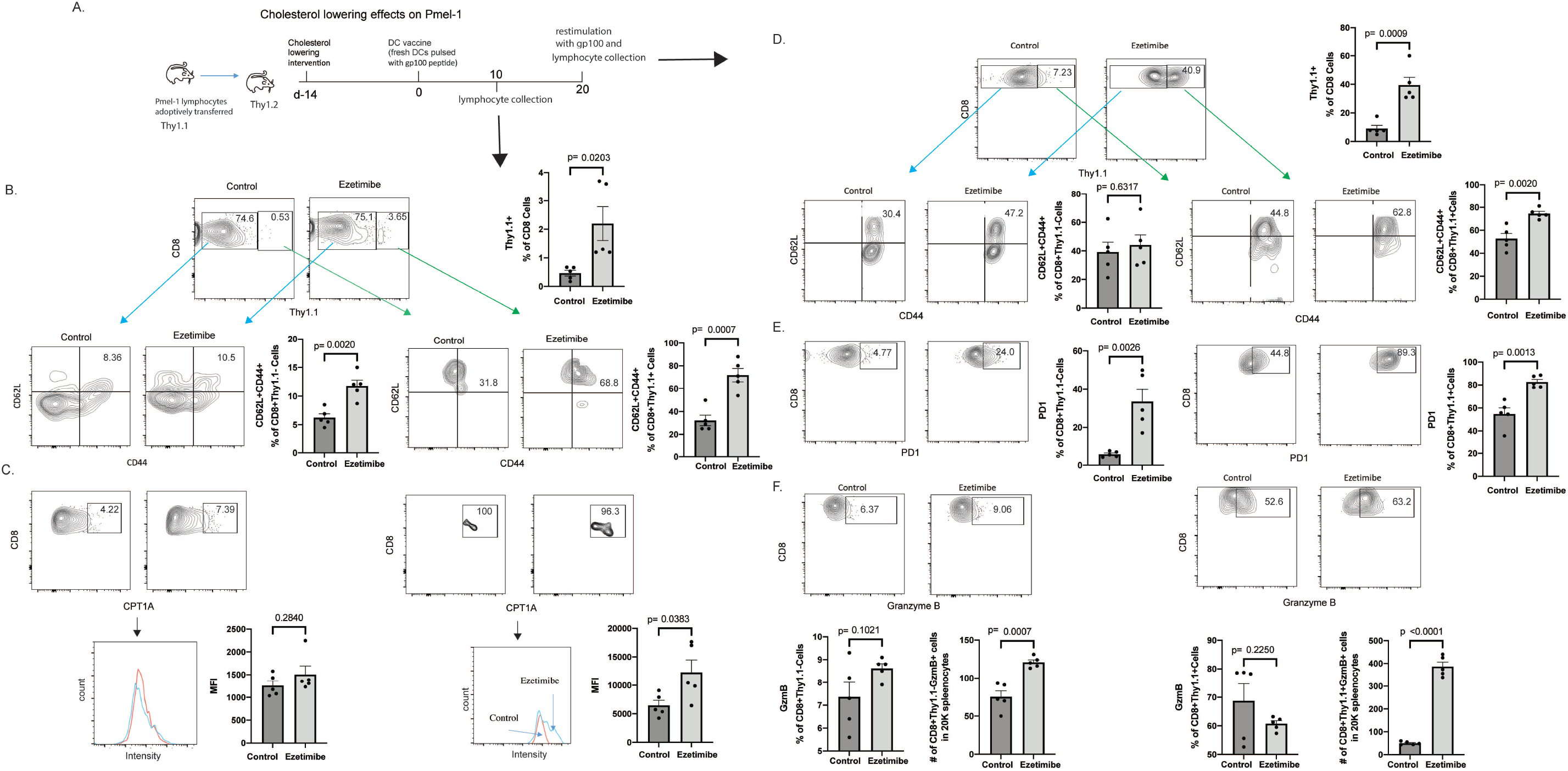
Cholesterol-lowering increases CD8+ T cell memory and enhances recalled response. **A**. Pmel-1 lymphocytes were adoptive transferred into congenic mice followed by ezetimibe treatment. Mice were injected with DCs pulsed with gp100 peptide on day 0 and day 20 (DC vaccination). Representative results are shown for triplicate experiments with 4-6 mice per experimental group. **B-C**. Lymphocytes were harvest on day 10 and analyzed for the indicated markers by flow cytometry. **B**. Donor Pmel-1 lymphocytes (Thy1.1+) were increased in ezetimibe-treated animals following DC vaccination and all CD8+ lymphocytes displayed a memory phenotype with increase in CD62L and CD44 expressing. **C**. CPT1A staining intensity was assessed in CD8+Thy1.1+ and CD8+Thy1.1-cells. **D-F**. Mice were restimulated with DC vaccination on day 20 and lymphocytes were harvested 3 days later. **D**. Ezetimibe treatment was associated with a greater recall proliferation of Pmel-1 CD8+ lymphocytes, which displayed a memory phenotype with increase in CD62L and CD44 expression. **E-F**. PD1 and Granzyme B expressions were assessd in CD8+Thy1.1+ and CD8+Thy1.1-cells.

**Figure 4.**
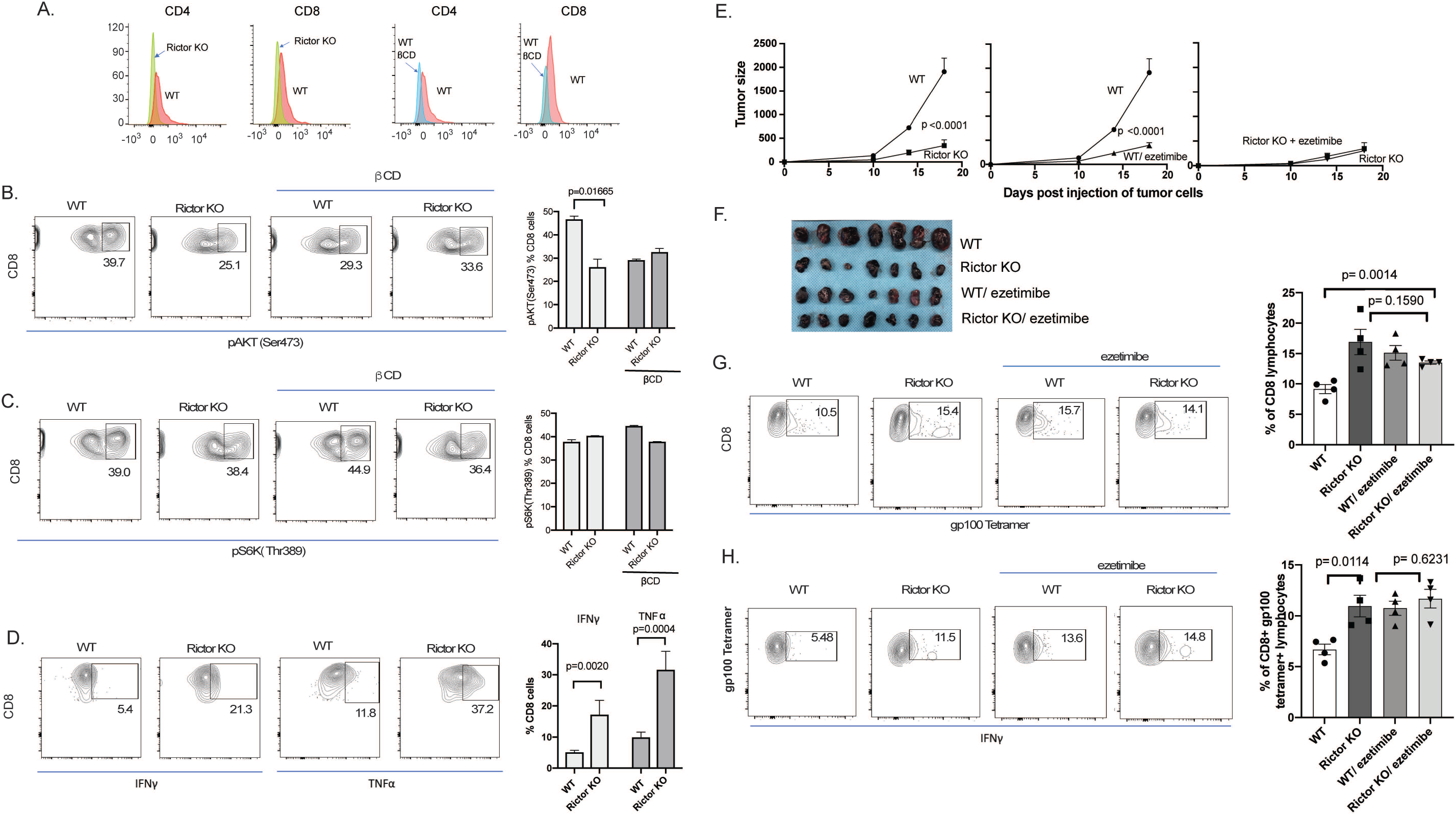
TORC2 knock out in lymphocytes enhances antitumor immunity. **A**. CD4+ and CD8+ lymphocytes from Rictor KO mice (left 2 panels) or wild type (WT; right 2 panels) were assessed for Rictor expression. **B**. CD8+ Lymphocytes from Rictor KO mice were assessed for pAKT (Ser473) by flow cytometry. β CD was added to cell culture to bind and reduce cholesterol in the medium. **C**. CD8+ lymphocytes were assessed for pS6k (Thr389). **D**. CD8+ lymphocytes were also stimulated with αCD3 and αCD28 to assess activation as measured by TNFα and IFN*γ* using flow cytometry. Summarized data (B-C) are for triplicate experiments. **E**. B16 tumors were also grown subcutaneously in mice with lymphocytes-specific Rictor KO and control (*Rictor*^flox^), with or without daily ezetimibe treatment. Tumors and spleens were harvested on day 18. Tumor growth was monitored and p-value was obtained by using the repeated measures ANOVA. **F**. Tumors were photographed. **G**. Splenocytes were stained with αCD8 and gp100 tetramer. **I**. Splenocytes were cultured *ex vivo* with gp100 peptide stimulation for 5 days. Lymphocytes were stained with αCD8, gp100 tetramer, and αIFN*γ* before flow cytometry analysis.

**Figure 5.**
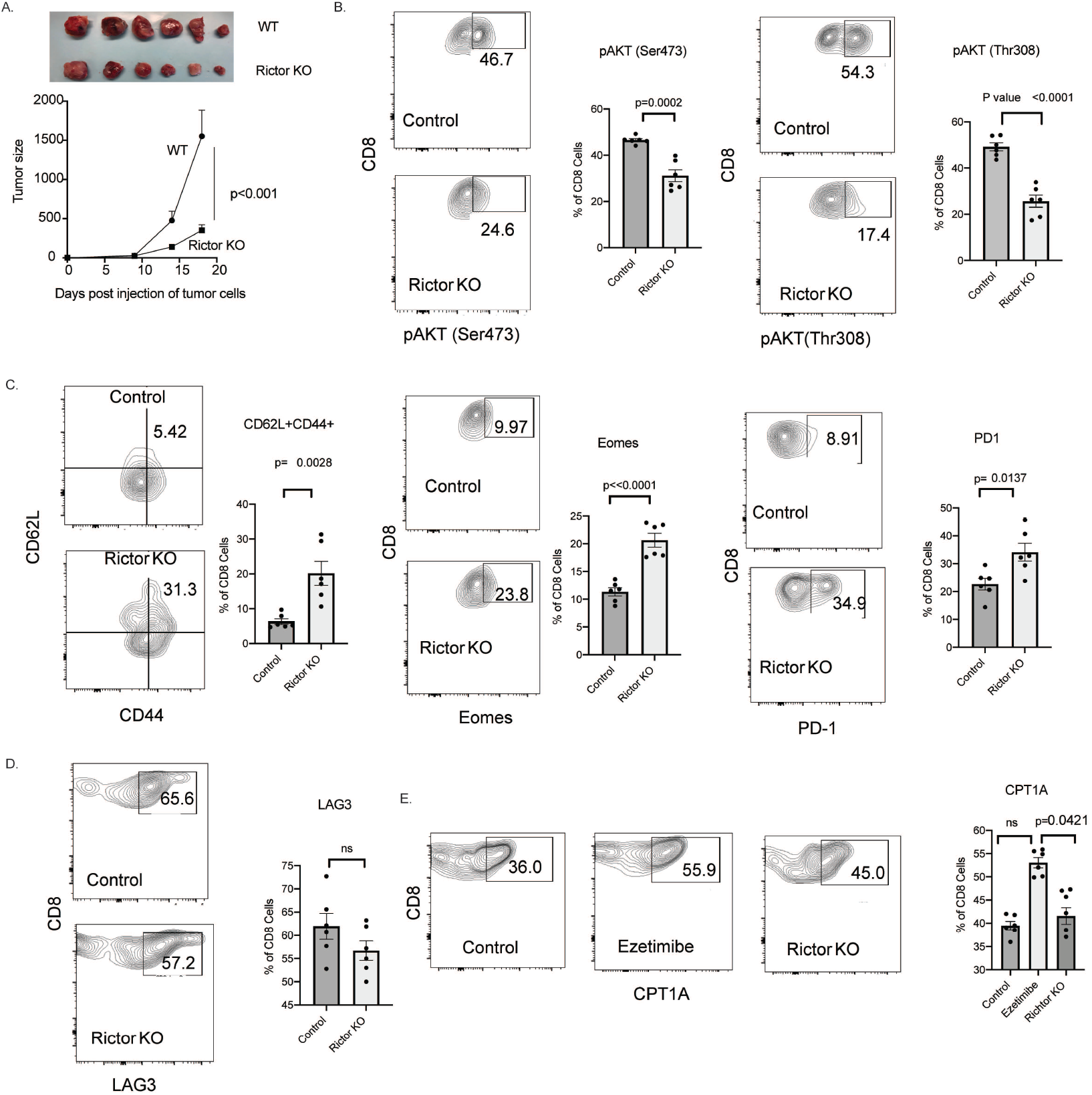
TORC2 knock out in lymphocytes increases markers for central CD8+ T cell memory and fatty acid oxidation. **A**. RM1 tumors were grown subcutaneously in mice with lymphocyte-specific Rictor KO (n=9) and control (*Rictor*^flox^, n=6). Tumor growth was monitored and on day 18, tumors were removed and photographed. Tumor growth was monitored and p-value was obtained by using the repeated measures ANOVA. **B-E**. Lymphocytes were collected from mice and analyzed for the indicated markers by flow cytometry. **G**. Splenocytes were first gated for CD8+ prior to flow cytometry analysis for the indicated markers.

The most important function of CD8+ memory cells is to expand rapidly when rechallenged. When Thy1.1+ lymphocytes were rechallenged with DCs pulsed with gp100 peptide on day 20, greatest expansion occurred in lymphocytes previously exposed to ezetimibe where approximately 40% of all CD8+ cells were Thy1.1+ lymphocytes **(Fig. 3D)**. The CD8+ lymphocytes maintained a central memory phenotype on day 23; they expressed CD62L and CD44. With CL intervention, both Thy1.1+ and Thy1.1-CD8 cells had increased PD1 expression, which has been shown to be required for both development and function of CD8+ memory T cells **(Fig. 3E)**.^29^ Finally, consistent with the increased CD8+ lymphocyte expansion upon stimulation after CL, both Thy1.1+ and Thy1.1-lymphocytes had an activated phenotype, expressing granzyme B, IFN*γ*, IL9 or T-bet after CL **(Fig. 3F, S. Fig. 5**,**6)**. These results suggested that lymphocytes stimulated by tumor in the presence of lower serum cholesterol exhibit enhanced CD8+ memory lymphocytes with a central memory phenotype.

### mTORC2 knock out in lymphocytes enhances antitumor immunity

A genetic approach was used to confirm that TORC2 is the target for CL that leads to enhanced antitumor immunity. Mechanistic target of rapamycin (mTOR) integrates cellular signals from nutrients, growth factors, hormones, stresses, and hypoxia. MTOR can complex either with raptor to form mTORC1 or Rictor to form mTORC2. Conditional knockout (KO) mice for mTORC2 were generated by crossing B6.Cg-Tg(Lck-cre)548Jxm/J with *Rictor*^*tm1*.*1Klg*^/SjmJ. Resulting mice had T lymphocyte-specific Rictor KO, Lck-Cre-*Rictor*^flox^ (Rictor-KO; **Fig. 4A, S. Fig. 7**). As expected, Rictor KO mice had decreased pAKT (Ser473), which is a target of TORC2 **(Fig. 4B)**. However, CL decreased Rictor protein expression **(S. Fig. 8, 9)**, which was unexpected and suggested this as the mechanism for decreased mTORC2 signaling for CL. Unlike the control lymphocytes, Rictor-KO lymphocytes did not respond to β-cyclodextrin (β-CD), which was added to lower cholesterol in the medium; CL did not enhance lymphocyte function via additional mechanisms beyond decreased Rictor. As expected, pS6K (Thr389), which is a target for mTORC1, was unchanged by KO of Rictor or β-CD **(Fig. 4C)**. Rictor KO lymphocytes had enhanced activation as indicated by TNFαor IFN*γ* expression in response to αCD3 and αCD28 **(Fig. 4D)**.

**Figure 7.**
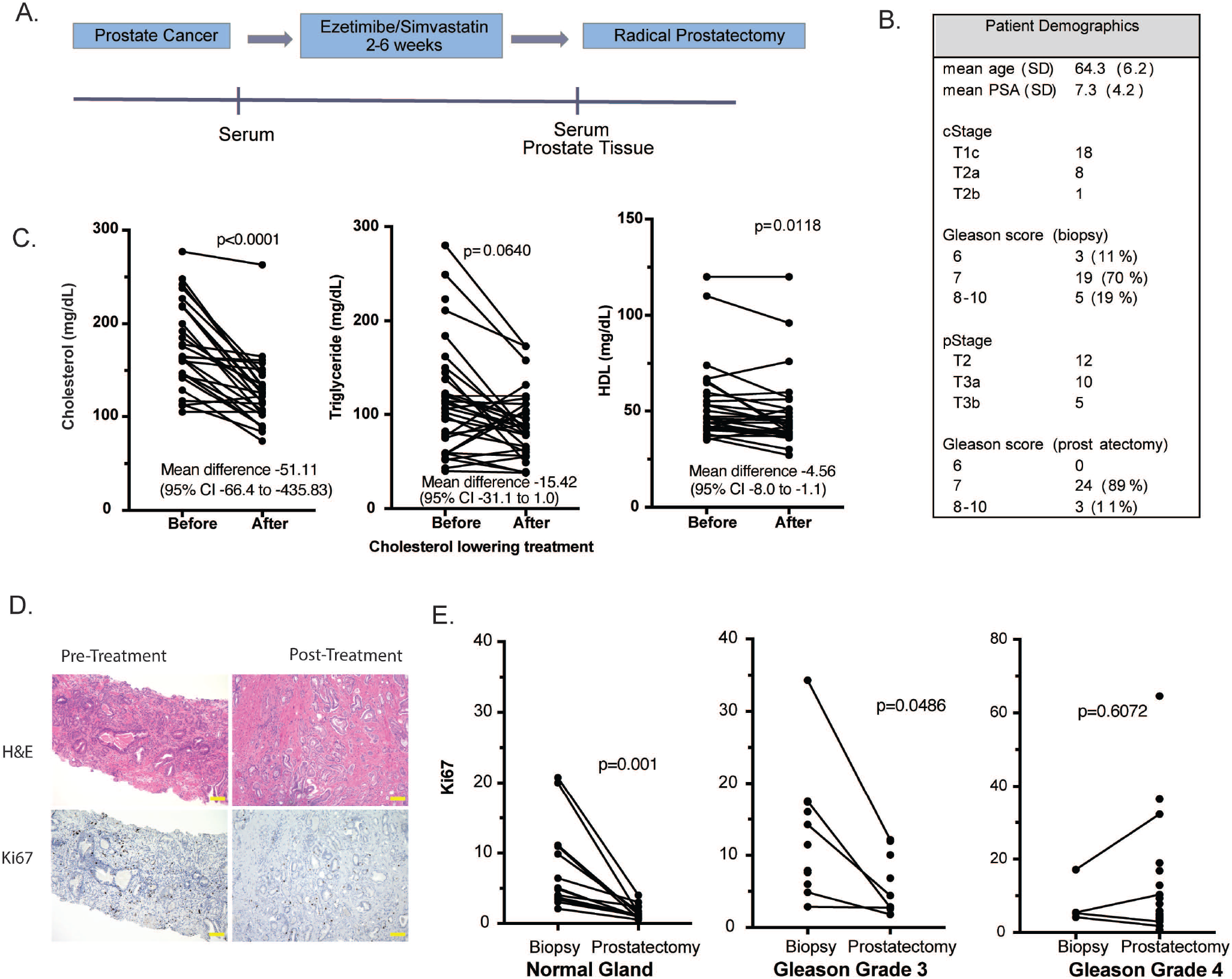
In patients, cholesterol lowering (CL) decreased cellular proliferation in the prostate. **F**. A prospective window-of-opportunity clinical trial was conducted where 27 men scheduled to undergo radical prostatectomy were treated preoperative with CL intervention. **G**. Serum lipids were measured before and at least 2 weeks after starting treatment. **H**. Ki67 immunohistochemical staining was quantified using pretreatment prostate biopsy tissue and posttreatment radical prostatectomy tissue. Scale bars represent 100μm **I**. Ki67 staining in normal prostate glands, Gleason grade 3 tumors, and Gleason grade 4 tumors were quantified. A line connects matching pre- and posttreatment tissue from the same patient.

To confirm *in vivo* that decreased signaling through mTORC2 in lymphocytes results in improved antitumor immunity, B16 or RM1 tumors were grown in Rictor-KO and control mice (*Rictor*^flox^). B16 tumor growth was suppressed in Rictor-KO mice compared to control mice (*Rictor*^flox^), and tumor suppression was similar to that with CL **(Fig. 4E, F)**. Both Rictor-KO and CL provided similar increase in tumor gp100-specific CD8+ lymphocytes **(Fig. 4G)**, which had increased IFN*γ* responses compared to lymphocytes from WT mice **(Fig. 4H)**. In Rictor-KO mice, CL provided no additional suppression of tumor growth, increase in gp100-specific CD8+ lymphocytes, or increase in gp100-specific IFN*γ* response **(Fig. 4E-H)**. These observations indicate that CL works on lymphocytes solely through suppression of Rictor and mTORC2.

To confirm these observations in another model, growth of RM1 tumors were compared in Rictor-KO and control mice. Tumor growth was significantly slower in Rictor-KO mice **(Fig 5A, S. Fig. 10)**. The CD8+ lymphocytes from Ricktor-KO mice had decreased mTOR2 signaling as indicated by decrease in pAKT (Ser473) and pAKT (Thr308) **(Fig 5B)**. The lymphocytes from Rictor KO mice had increased expression of Granzyme B and IFN*γ* **(S. Fig. 11)**. The Ricktor-KO mice had increased in markers of central memory, CD8+ cells with increased CD62+CD44+ cells, CD127+KLRG11-cells, Eomes, and PD1 **(Fig. 5C, S. Fig. 12)**. However, LAG3, which is a marker of T cell exhaustion was not increased by Rictor KO **(Fig. 5D)**. Fatty acid oxidation (FAO) has been linked to increased central T cell memory formation. The rate-limiting step of fatty acid oxidation is mediation by CPT1A. CPT1A expression increased with CL but not with Rictor KO **(Fig. 5E)**. This suggests that CL decreases FAO, which in-turn decreases mTOR2 signaling.

The mTOR/TORC1 pathway has been shown to regulate CD8+ T cell memory formation.^24^ Rapamycin and rapamycin analogs, which inhibit TORC1, have been proposed as immunotherapy to enhance CD8+ T cell memory formation.^23^ However, serum CL had the greatest effect on signaling through the TORC2 pathway. Lymphocyte-specific TORC2 knockouts demonstrated enhanced lymphocyte function and antitumor capacity both *in vitro* and *in vivo*, in multiple models. This is in line with a report by Pollizzi et al., who examined the role of mTORC1 and mTORC2 on CD8+ T cell differentiation.^30^ They found that mTORC1 is critical to CD8+ T cell activation; however, mTORC2 regulates CD8+ T cell memory. No mTORC2-specific inhibitors are available for clinical testing but we describe a clinically available intervention to specifically inhibit mTORC2 signaling, which may work by suppressing Rictor expression. Therefore, mTORC2 inhibition appears both safe and clinically feasible using approved CL therapies.

### CL increases CD8+ T cells in tumor and decreased tumor proliferation

Another consequence of enhanced antitumor immunity is increased infiltration of lymphocytes into the tumor. Our animal studies revealed that CL increased CD8+ lymphocytes in prostate tumor **(Fig. 6A)**. In an exploratory human study, RNAseq was performed to quantify immune cells present in prostate cancer **(Fig. 6B)**. Two patients were examined, and both showed increased CD8+ lymphocytes and M1 macrophages following CL intervention with the combination of ezetimibe and simvastatin. This observation was confirmed in six additional patients scheduled to undergo radical prostatectomy who were treated with 2-6 weeks of CL. Matched pre-treatment prostate biopsy and post-treatment prostatectomy tissue showed that CL therapy increased CD8+ lymphocytes in the tumor **(Fig. 6C)**.

**Figure 6.**
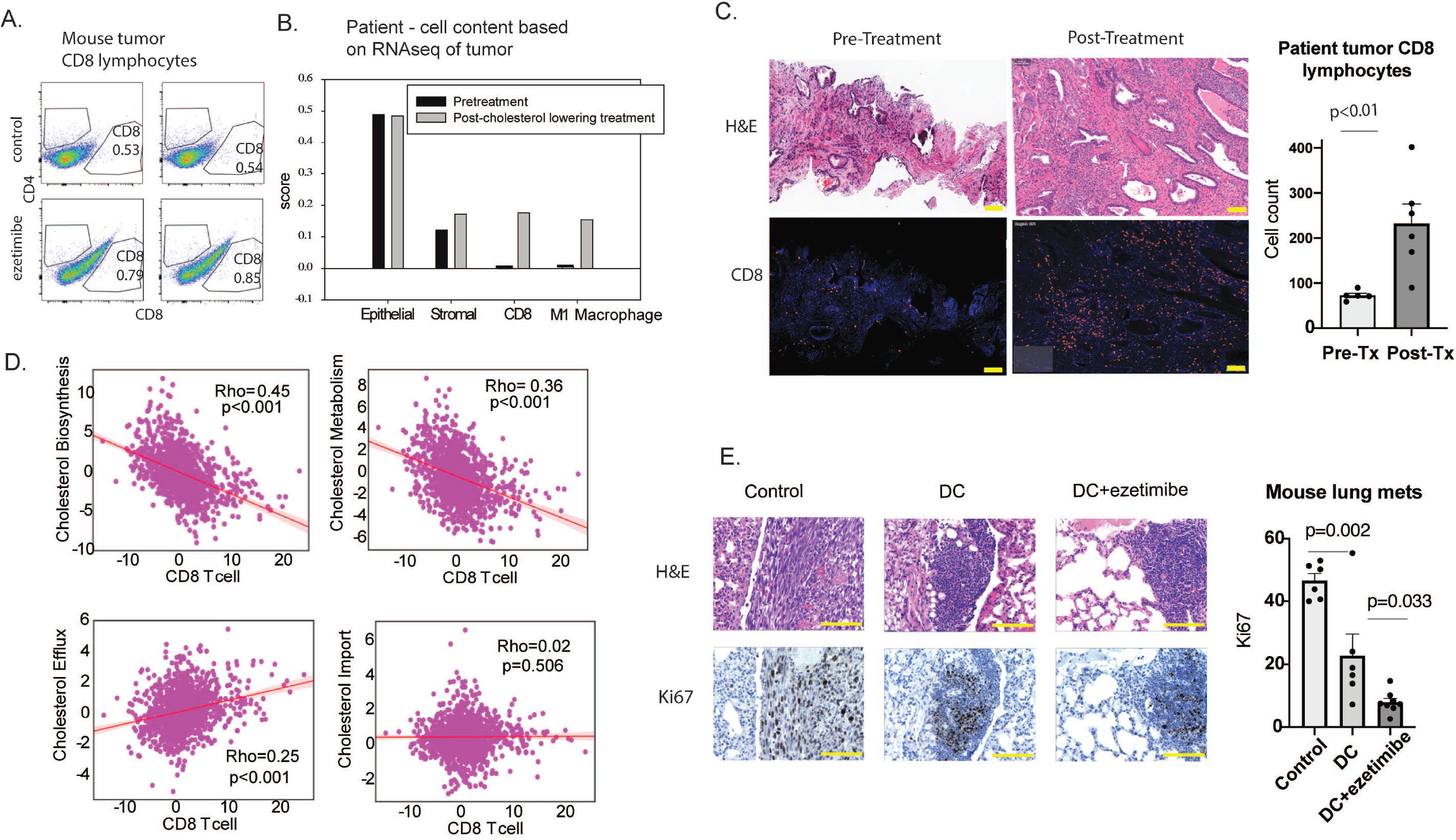
Cholesterol lowering (CL) increased infiltration of CD8+ lymphocytes into tumor (mice and human) and decreased tumor proliferation (mice). **A**. CL with ezetimibe increased tumor infiltration of CD8+ lymphocytes in mice (p=0.03). Mouse experiments were performed in duplicate and representative result is shown. **B**. In patient tumor tissue, CL with a statin and ezetimibe increased tumor CD8+ lymphocytes and M1 macrophages as determined by RNAseq. This analysis was performed for 2 patients and a representative result is shown. **C**. In similarly treated patients, αCD8 immunofluorescent staining are shown using matched pre-treatment prostate biopsy and post-treatment prostatectomy tissue. For the summarized data, each column shows mean values from 6 matched pre-treatment biopsies and post-treatment prostatectomies. **D**. Using gene expressions from 1,321 tumors in the PCTA database, scatter plot and regression line were generated to show the relationship between genes involved in cholesterol homeostasis (biosynthesis, metabolism, efflux and import) and genes specific for CD8+ T cells. The correlation coefficient was computed by Spearman’s method. **E**. Using the RM1 lung metastases from the experiment described in Figure 1H, representation H&E section and Ki67 staining are shown. All scale bars represent 100μm (C,E).

To provide additional evidence of a relationship between cholesterol and CD8+ lymphocytes, we analyzed gene expression from the Prostate Cancer Transcriptome Atlas, which is a virtually organized database of 1,321 clinical specimens from 38 prostate cancer cohorts **(Fig. 6D)**.^31^ There was a strong, significant inverse correlation between genes specific for CD8+ T cells and genes involved in cholesterol biosynthesis and metabolism. There was a significant positive correlation between genes specific for CD8+ T cells and genes involved in cholesterol efflux, which would be involved in removing intracellular cholesterol, but not with genes involved in cholesterol import. Taken together, these results suggest that low cholesterol increases CD8+ lymphocyte infiltration in the tumor.

If CL produces antitumor immunity, the end result is expected to be a decrease in tumor proliferation. In the murine experiment described in **Fig. 1H**, lymphocytes from mice treated with a DC vaccine and CL were transferred to na^ï^ve mice challenged with IV tumor cells. The resulting decrease in cellular proliferation measured using Ki-67 staining of lung metastases was due to the transferred lymphocytes since the tumor bearing mice were never exposed to the DC vaccine or CL intervention **(Fig. 6E)**.

### CL decreased proliferation in patient prostates

To see if CL decreases Ki-67 in human prostate cancers, we used archival prostate tissue from a previously conducted, prospective clinical trial where men received 2-6 weeks of an aggressive cholesterol lowering intervention prior to radical prostatectomy **(Fig. 7A)**. The study targeted two sources for serum cholesterol in humans: simvastatin inhibited cholesterol synthesis in the liver and ezetimibe decreased cholesterol absorption in the intestine. Patient characteristics are summarized in **Fig. 7B**. There was a large, significant decrease in serum cholesterol and modest decrease in triglyceride (nonsignificant) and HDL on the trial **(Fig. 7C)**. When comparing pre and post-treatment prostate tissue, apoptosis assessed using Caspase 3 staining did not change (data not shown). However, proliferation decreased in normal prostate tissue and low-grade prostate cancers **(Fig 7D,E)**. There was no significant change in Ki-67 staining in high-grade prostate cancers. These results suggest that currently available therapies for cholesterol lowering may decrease growth in both benign prostate that produces voiding symptoms in older men and low-grade prostate cancer, which is managed with active surveillance because of the risk of progression to aggressive cancer.

## DISCUSSION

The mTOR/TORC1 pathway has been shown to regulate CD8+ T cell memory formation.^24^ In fact, rapamycin and rapamycin analogs, which inhibit TORC1, have been proposed as immunotherapy that can enhance CD8+ T cell memory formation.^23^ Therefore we assessed the mTOR pathway in lymphocytes from mice treated with ezetimibe. We were surprised to find that lowering serum cholesterol had the greatest effect on signaling through the TORC2 pathway. Therefore, lymphocyte-specific TORC2 knockouts were generated and shown to have enhanced lymphocyte function and antitumor capacity. It is not clear if FAO is necessary for CD8+ T cell memory formation^32, 33^, however, the rate-limiting step of FAO mediated by CPT1A has been associated with CD8+ T cell memory formation^26^. We found that increased CPT1A was associated with CL but not with Ricktor KO, suggesting CL first increases FAO, which then inhibits mTOR. Both CL and TORC2 knockout increased central memory CD8+ lymphocytes. This is in line with a report by Pollizzi et al., who examined the role of mTORC1 and mTORC2 on CD8+ T cell differentiation.^30^ They found that mTORC1 is critical to CD8+ T cell activation; however, mTORC2 regulates CD8+ T cell memory. There are currently no mTORC2-specific inhibitors in clinical testing and we now describe the only clinical intervention to specifically inhibit mTORC2, and the intervention is safe and already clinically available.

We confirm that our observations extend to patients. We used archival tissue from a previously conducted, prospective window-of-opportunity clinical trial where patients scheduled to undergo prostatectomy for prostate cancer were treated with 2-6 weeks of both ezetimibe and simvastatin to achieve maximal CL. In mice we reduced cholesterol by approximately 10% to see the immune response. In patients we achieved a greater CL, which was approximately 25% of baseline. We showed that post-treatment lymphocytes had a decrease in mTORC2 signaling. In our mouse model, CL enhanced CD8+ T cell functions and increased CD8+ T cells in the tumor. Using RNAseq and immunofluorescence in clinical prostate cancers, we showed that post-treatment tumors exhibited an increase in CD8+ T cells when compared to matched pre-treatment tumors. Finally, we used a large, publicly available prostate cancer transcriptome dataset with over 1000 patient prostate cancers to show an inverse relationship between CD8+ lymphocyte infiltration and genes involved in cholesterol biosynthesis and metabolism. Also consistent with the observation that lower cholesterol in the tumor promotes CD8 antitumor immunity, there was a positive correlation between CD8+ T cell infiltration and cholesterol efflux genes.

These results suggest that clinical interventions to lower serum cholesterol can stimulate antitumor immunity by enhancing CD8+ memory formation. Localized prostate cancer is a slow growing cancer where only a small minority of patients will eventually die from their disease, even when treatment is not provided. Therefore, any therapy targeting this population should have minimal morbidity. In the case of statins, not only is the therapy safe but there are other potential cardiovascular health benefits, further strengthening the case for CL intervention. Our results provide justification for formal clinical testing of CL intervention for primary or secondary chemoprevention and as a strategy to enhance the effects of immunotherapies for advanced solid tumors.

## Conclusion

In clinically relevant murine models for assessing antitumor immunity, we show that CL inhibited tumor growth by enhancing CD8+ lymphocytes, which have enhanced memory function and infiltrated tumor tissue. Serum CL inhibited mTOR2 signaling in mouse lymphocytes. We confirmed that our observations extend to patients by examining lymphocytes and prostate tissue from a prospective window-of-opportunity clinical trial where patients scheduled to undergo prostatectomy for prostate cancer were treated with 2-6 weeks of ezetimibe and simvastatin to achieve maximal CL.

## METHODS

### Antibodies and reagents

The following monoclonal antibodies (mAb) for flow cytometry, with or without a fluorescent conjugate, were obtained from Biolegend (San Diego, CA): αCD4 (GK 1.5 and RM4-5), αCD8 (53-6.7), αT-bet (4B10), αCD62L (MEL-14), αFoxP3 (FJK-16s), αIFN-*γ* (XMG1.2), αTNFα (TN3-19.12). αCD4 (GK1.5) and αCD8 (2.43) antibodies for T cell depletion were purchased from BioXcell (West Lebanon, NH). Ezetimibe was purchased from Par Phamaceutical (Chestnut Ridge,NY). Antibodies for Western blotting against the following targets were purchased from Cell Signaling Technology (Danvers, MA): P-p70S6K (T389), p70 S6K,P-AKT (S473), AKT(C67E7), P-4E-BP1 (T37/46), 4E-BP1 (53H1), P-AKT (T308). Antibody against β-actin (AC-15) was purchased from Sigma (Ronkonkoma, NY).

### Western blot

Cells were washed with ice-cold PBS and lysed with RIPA buffer (50 mM Tris-HCl pH 7.5, 150 mM NaCl, 1 mM EDTA, 10 mM DL-Dithiothreitol, 10% glycerol, 0.5% sodium deoxycholate, 1% Triton-X100) with 10 mM NaF, protease inhibitors (1:100 dilution of protease inhibitor cocktail from Sigma). Protein concentrations were measured with the Bio-Rad Protein Assay (Hercules, CA). 40 μg of lysate was loaded for Western blot analysis. Nitrocellulose membranes were used for the protein transfer and western blots were performed according to recommendations of the antibody manufacturer.

### Tumor cells

RM1, RENCA and B16 cancer cell lines were purchased from ATCC (Manassas, Virginia). The cells were maintained in Dulbecco’s modified Eagle medium or RPMI 1640 medium supplemented with 10% heat-inactivated FBS (Gemcell, West Sacramento, CA), 2 mmol/L of L-glutamine, 100 units/mL of penicillin, 100 μg/mL of streptomycin, and 250 ng/mL of Amphotericin B (ThermoFisher, Waltham, MA). All cells were periodically authenticated by morphologic and histologic inspection and animal grafting for assessing their ability to grow and metastasize. The cells were annually tested for mycoplasma using Myco Alert Kit (Lonza, Allendale NJ).

### Mice

C57BL/6J, Balb/C and Pmel-1 mice, 5-8 weeks old were purchased from Jackson laboratory (Ellsworth, Maine) and housed under pathogen free conditions. All experiments involving animals were in compliance with federal and state standards, which include the federal Animal Welfare Act and the NIH guide for the care and use of laboratory animals. Tumors were generated by subcutaneously injecting 2×10^5^ tumor cells into the frank. To lower serum cholesterol, mice were administrated daily ezetimibe (30mg/Kg) by oral gavage for 2 weeks before injection of tumor cells and continuing throughout the experiment. To deplete CD8+ lymphocytes, mice were injected intraperitoneally with 200 μg of CD8 mAb (clone 2.43). Tumor size was measured with calipers approximately twice a week and tumor volume was calculated (shortest diameter^2^ x longest diameter/2). Blood was drawn from the tail vein, and the serum cholesterol concentration was determined using Amplex^™^ Red Cholesterol Assay Kit (Invitrogen, Carlsbad, CA). For the lung metastasis model, 2×10^5^ B16 or RM1 cells were intravenously injected through the tail vein. The lung tissues were harvested to determine lung weight and count tumor deposits using a dissection microscope. To monitor CD8+ T cell memory formation *in vivo*, Pmel-1 lymphocytes were harvested from Thy1.1+ mice and 2×10^6^ lymphocytes were adoptively transferred by tail vein injection into Thy1.2+ mice. For all syngeneic tumor studies in mice, each experimental group had 4-10 mice.

Conditional knockout (KO) mice for mTORC2 were generated by crossing B6.Cg-Tg(Lck-cre)548Jxm/J with *Rictor*^*tm1*.*1Klg*^/SjmJ (both from Jackson labs). Resulting mice had lymphocyte-specific Rictor KO, *Lck-Cre-Rictor*^flox^ (Rictor-KO). Homozygous mice were obtained by interbreeding heterozygotes. Genotyping was performed by PCR. The primer sequences were: Rictor forward, CAA GCA TCA TGC AGC TCT TC; Rictor reverse TCC CAG AAT TTC CAG GCT TA; Cre forward, TGT GAA CTT GGT GCT TGA GG; Cre reverse, CAG GTT CTT GCG AAC CTC AT.

### Dendritic cell vaccine and T-cell stimulation

To prepare dendritic cell (DC) vaccine, bone marrow was harvested from C57BL/6 mice and suspended in RPMI supplemented with 10% FBS. GM-CSF (10 ng/mL) was added to the medium and placed at 37°C with 5% CO2 for 7 days. DCs were pulsed with tumor cell lysate for 16 hours and activated with 10 μg/mL CpG for 4 hours. Alternatively, DCs were pulsed with 10 ng/mL mouse gp100 peptide (amino acids 25–33, which is presented by H2-Db class I molecules; Alpha Diagnostic International, San Antonio, TX) for 1 hr and activated with 10 μg/mL CpG for 2 hours. The resulting DC vaccine was injected subcutaneously into mice. For *in vitro* activation of Pmel-1 cells, DCs were washed with PBS and cocultured with Pmel-1 cells, which were harvested from spleen of Pmel-1 mice. IFNγ, IL-9 and TNFα were analyzed by flow cytometry (LSR II, BD Biosciences, San Jose, CA 95131).

### Adoptive transfer protocol

To monitor CD8+ T cell memory *in vivo*, spleens and lymph nodes were harvested from Thy1.1+ Pmel-1 mice. The spleen and lymph nodes were transferred to a 70 micron cell strainer on a 50 mL tube. A 1 mL syringe plunger was used to gently press the tissue through the strainer using 10 ml red cells lysis buffer to wash the strainer. Cells were then spun at 1500rpm for 5 min to collect dissociated lymphocytes. CD8+ cells were isolated using the Mouse CD8 Cell Recovery Column Kit from Cedarlane Laboratories Ltd (ON, Canada). 1×10^5^ CD8+ enriched cells were stained with CD8-PE-Cy5 and thy1.1-FITC antibodies to confirm presence of CD8 and Thy1.1 by flow cytometry. 2×10^6^ Pmel-1 lymphocytes were adoptively transferred by tail vein injection into Thy1.2+ C57BL/6 mice.

### Isolation of tumor infiltrating lymphocytes

Mouse tumors were cut into small fragments ∼2–3 mm in length and put in an enzyme digest mix consisting of 10 μg/mL DNAse, 1 mg/mL collagenase and 25μg/mL hyaluronidase (all from Sigma) and incubated for 3 hours at 37° C under continuous rotation. Tumor infiltrating lymphocytes (TILs) were isolated by Ficoll-Paque density gradient centrifugation. CD4 and CD8 cells were analyzed by flow cytometry (LSR II).

### Immunohistochemistry

Patient prostate tissues or mouse tumors were fixed in 10% formalin in PBS and embedded in paraffin. 4μm sections were baked at 60°C for 1 hr and subjected to Immunofluorescence staining using the Ventana Discovery Ultra autostainer with buffers and antibodies from Ventana Medical Systems (Oro Valley, AZ). Briefly slides were deparaffinized using EZ solution, followed by antigen retrieval with Cell Conditioning 1 (CC1) solution (prediluted Tris solution, pH 8.0) for 64 min at 95°C and blocking with Discovery inhibitor (Ventana) for 12 min at RT. The slides were incubated with αCD8 (Ventana) or αKi-67 (Ventana) at 37°C for 28 minutes followed by detection with anti-rabbit-HQ/ αHQ-HRP/ DAB (Ventana) or Red610 kits (Ventana). DNA dye DAPI (Invitrogen) was used to stain the nucleus. Slides was scanned using TissueFAXS 200 system (TissueGnostics, Tarzana, CA). Image was analyzed using QuPath (Edinburgh, UK) with default settings. For all summarized data, 3 random high-power fields were evaluated by automated image analysis and averaged prior to generating a mean for each experimental group.

### Clinical trial

A prospective, window-of-opportunity trial was previously conducted (ClinicaltTrials.gov identifier: NCT02534376) and provided tissue and serum. Patients with clinically significant prostate cancer scheduled to undergo prostatectomy provided written consent and were enrolled in Pro40945, which is a clinical trial that received ethics approval from the Cedars Sinai Medical Center’s Institutional Review Board (Los Angeles, CA). Patients received two-six weeks of maximal CL intervention with Vytorin, which contains ezetimibe (10mg) and simvastatin (40mg). Whole blood and prostate biopsy tissue were collected prior to starting Vytorin and posttreatment blood and prostate tissue were collected at the time of prostatectomy. Following radical prostatectomy, prostate specimens were fixed in 10% neutral buffered formalin and processed according to the International Society of Urological Pathology Consensus Guidelines on prostatectomy handling.^34^ Prior to any studies of human prostate tissue, the H&E sections were examined by an expert genitourinary pathologist to identify the dominant tumor and to annotate it for benign prostate, tumor and tumor grade. These annotations were used for immunohistochemistry image analysis and for microdissection of tissue for RNA extraction. Alliance of Clinical Trials in Oncology funded the clinical trial, and their publication committee reviewed and approved this manuscript prior to submission for publication.

### RNAseq and tumor microenvironment

Tumors were macrodissected for RNA extraction. Tumor sections were placed in 2.0 mL RNase-free Eppendorf tubes. Sections were treated twice with 1.9 mL xylene for 5 min at 55 °C while rocking. The sections were washed twice with 100% ethanol. RNA was extracted from the paraffin samples using the MasterPure^™^ RNA Purification Kit (Epicenter Biotechnologies, Madison, WI, USA). RNA was then treated with 20 units DNase I (Ambion, Austin, TX, USA) for 30 min and checked for residual genomic DNA by TaqMan RT-PCR targeting ACTB. The final RNA concentration (A260:0.025) and purity (A260:A280 ratio) were measured using a NanoDrop ND-2000 spectrophotometer (NanoDrop Technologies, Wilmington, DE, USA).

For RNA sequencing, 300 ng total RNA were sent to MedGenome Inc (Foster City, CA), where RNAseq was performed on the HiSeq-2500 platform (Illumina, San Diego, CA) after using the TruSeq RNA Access Library Prep Kit (Illumina). Reads mapping to ribosomal and mitochondrial genome were removed before performing alignment with STAR (2.4.1).^35^ Overall 96% of the total pre-processed reads mapped to the reference gene model/genome. The raw read counts were estimated using RSEM.^36^ Read count data were normalized by trimmed mean of M-values (TMM) method using DESeq2. Tumor microenvironment analysis was performed using ESTIMATE software^37^ for all the protein coding genes at multiple levels i.e. tumor content, innate immune cells and adaptive immune cells. CD8+ T cell signature was obtained from CIBERSORT^38^ and cholesterol biosynthesis signature was from the molecular signature database (MSigDB)^39^. Individual signatures were used to compute enrichment scores for CD8+ T cell infiltration and cholesterol in prostate cancer using the Prostate Cancer Transcriptome Atlas (http://www.thepcta.org). The correlation coefficient was computed by Spearman’s method. Enrichment scores for the signatures were computed using weighted z-score method^40^ with intrinsic functionality of the PCTA. Mean values and S.E.M. are provided throughout the manuscript and p-values were generated using t-tests. Mouse tumor growth curves were compared using the repeated measures ANOVA.

## Data Availability

The RNAseq data is available at GEO (accession number GSE159529.

## Data Availability

The RNAseq data is available at GEO (accession number GSE159529).

## Acknowledgements

This work was supported by grants from the Samuel Oschin Comprehensive Cancer Center, Winnick Family Foundation, Homer and Gloria Harvey Family Endowed Chair, Teshinsky Family Foundation and the National Institutes of Health (R01CA182438).

The clinical trial reported in this publication was supported by the National Cancer Institute of the National Institutes of Health under the Award Number UG1CA189823 to the Alliance for Clinical Trials in Oncology NCORP Research Base (Jan C. Buckner, M.D., contact PI). The content is solely the responsibility of the authors and does not necessarily represent the official views of the National Institutes of Health.

## Author Contributions

Conceptualization, Y.W., J.L.M. and H.L.K.; Methodology, Y.W., S.Y., A.Y., S.S., M.R.F., and H.L.K.; Investigation, Y.W., S.Y., A.Y., E.M.L., S.K. and S.S.; Writing – Original Draft, Y.W. and H.L.K.; Writing – Review & Editing, Y.W., S.Y., A.Y., S. S., M.R.F., E.M.L., J.L.M. and H.L.K.; Funding Acquisition, H.L.K.; Resources, H.L.K.; Supervision, Y.W. and H.L.K.

## Figure Legends

**S. Figure 1. Transfer of activated lymphocytes inhibits s.c. growth of RM1 tumors**. B6 mice were treated with or without ezetimibe for 14 days prior to immunization with DCs pulsed with RM1 lysate. Lymphocytes were transferred to na^ï^ve mice which were challenged with s.c. RM1 tumors. Tumor growth was assessed. Each experimental group contained 4 mice.

**S. Figure 2. Splenocytes from ezetimibe-treated mice showed decreased pAKT (Ser473) in CD8+ T cells**. Mice were treated with 14 days of ezetimibe and untreated controls were examined by flow cytometry. Each experimental group contained 5 mice and is representative of duplicate experiments.

**S. Figure 3. DC vaccine was prepared**. Bone marrow was harvested from C57BL/6 mice and suspended in RPMI with GM-CSF. DCs were pulsed with gp100 peptide and activated with CpG. The resulting DCs were characterized by flow cytometry.

**S. Figure 4. Cholesterol-lowering intervention enhances CD8+ cells**. Pmel-1 lymphocytes were adoptive transferred into congenic mice (n=6 per group) followed by ezetimibe treatment and DCs pulsed with gp100 peptide. This figure represents a replicate of the experiment in Figure 3, but with addition markers. Donor Pmel-1 lymphocytes were assessed 10 days following DC vaccination. Thy1.1 expressing donor cells were assessed by flow cytometry. Lymphocytes expressing CD8 were assessed by flow cytometry for the indicated markers.

**S. Figure 5. Cholesterol-lowering intervention enhances CD8+ memory phenotype**. Pmel-1 lymphocytes were adoptive transferred into congenic mice (n=6 per group) followed by ezetimibe treatment and DCs pulsed with gp100 peptide. This figure represents a replicate of the experiment in Figure 3. **Donor Pmel-1 lymphocytes were assessed 30 days following DC vaccination**. Cells were restimulated *ex vivo* with gp100 peptide before flow cytometry. **Thy1.1 expressing donor cells** were assessed by flow cytometry. Lymphocytes expressing CD8 were assessed by flow cytometry for the indicated markers.

**S. Figure 6. Cholesterol-lowering intervention increases the number of CD8+ cells expressing Granzyme B**. This experiment uses lymphocytes harvest on day 10 of the experiment in Figure 3. CD8+ lymphocytes with and without Thy1.1 expression were analyzed by flow cytometry for Granzyme B expression.

**S. Figure 7. Conditional Rictor KO mice were generated. Genotyping results are shown**. Lanes 1 and 2 show mice with *loxP* sites flanking exon 11 of *Rictor*. Mouse represented by Lane 1 also has Cre under the control of the *Lck* promoter (Rictor KO). The mouse represented by Lane 2 is the control mouse.

**S. Figure 8. Rictor expression was measured in lymphocytes from conditional Rictor KO mice**. Lymphocytes were gated on CD4 (left) or CD8 (right). Rictor expression was assessed by flow cytometry in Rictor KO lymphocytes, with or with βCD.

**S. Figure 9. Rictor expression was measured in experimental replicates**. Experimental replicates (n=4) are shown for experiments shown in Figure 5A and S. Figure 7.

**S. Figure 10. B16 tumors were harvest and weighted**. In the experiment described in Figure 5A, mouse tumors were harvested 18 days following tumor cell injection. The tumors in each experimental group were weighted and averaged.

**S. Figure 11. Granzyme B and IFNγ were measured in lymphocytes from conditional Ricktor KO mice**. Flow cytometry was used to measure the indicated markers.

**S. Figure 12. CD127+KLRG1-lymphocytes from conditional Ricktor KO mice were quantified**. Flow cytometry was used to measure the indicated markers after gating on CD8.

