## Supplemental figures for "Cholesterol-lowering Decreased mTOR Complex 2 Signaling and Enhanced Antitumor Immunity"

### Slide 1
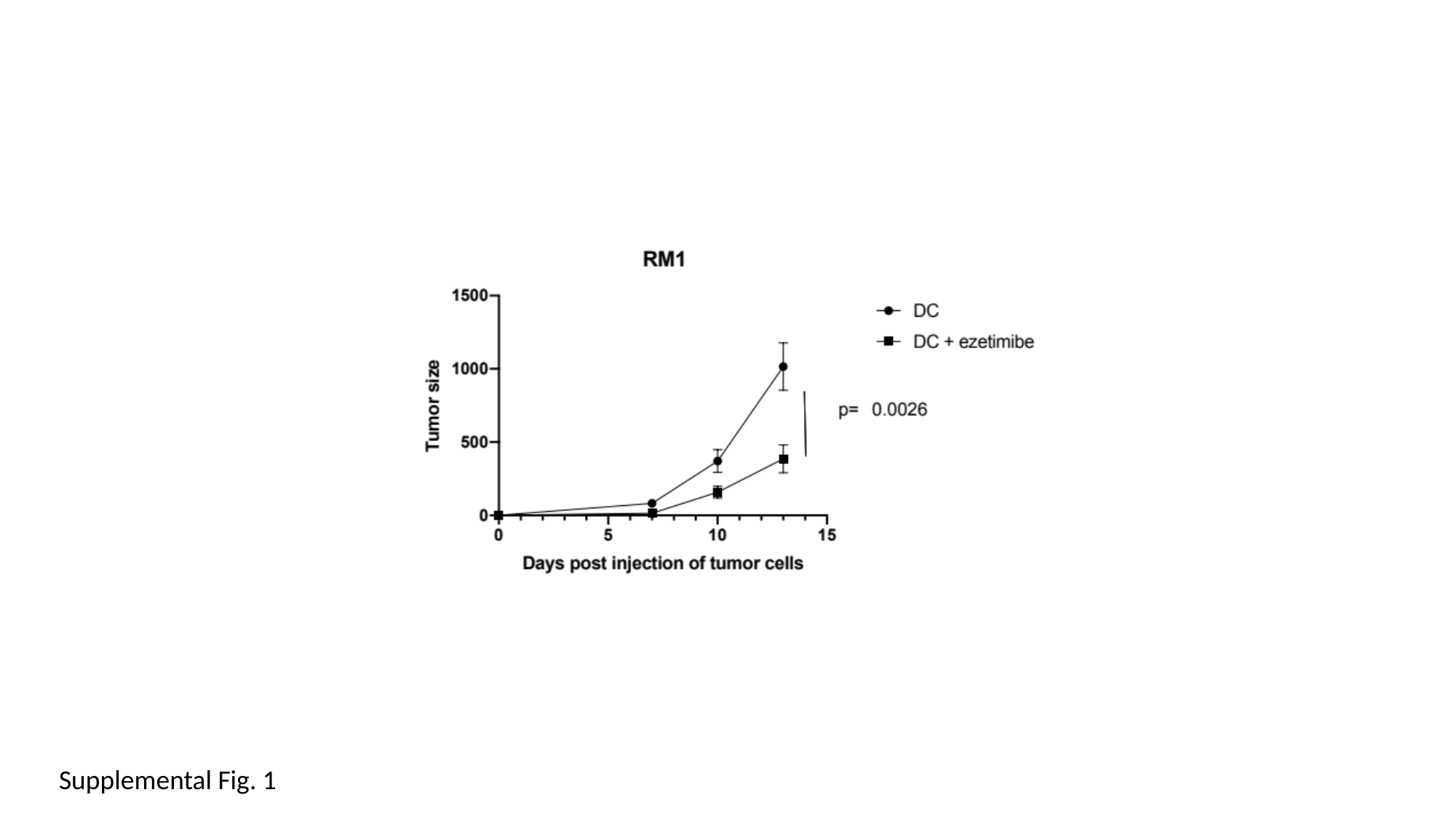

Supplemental Fig. 1

### Slide 2
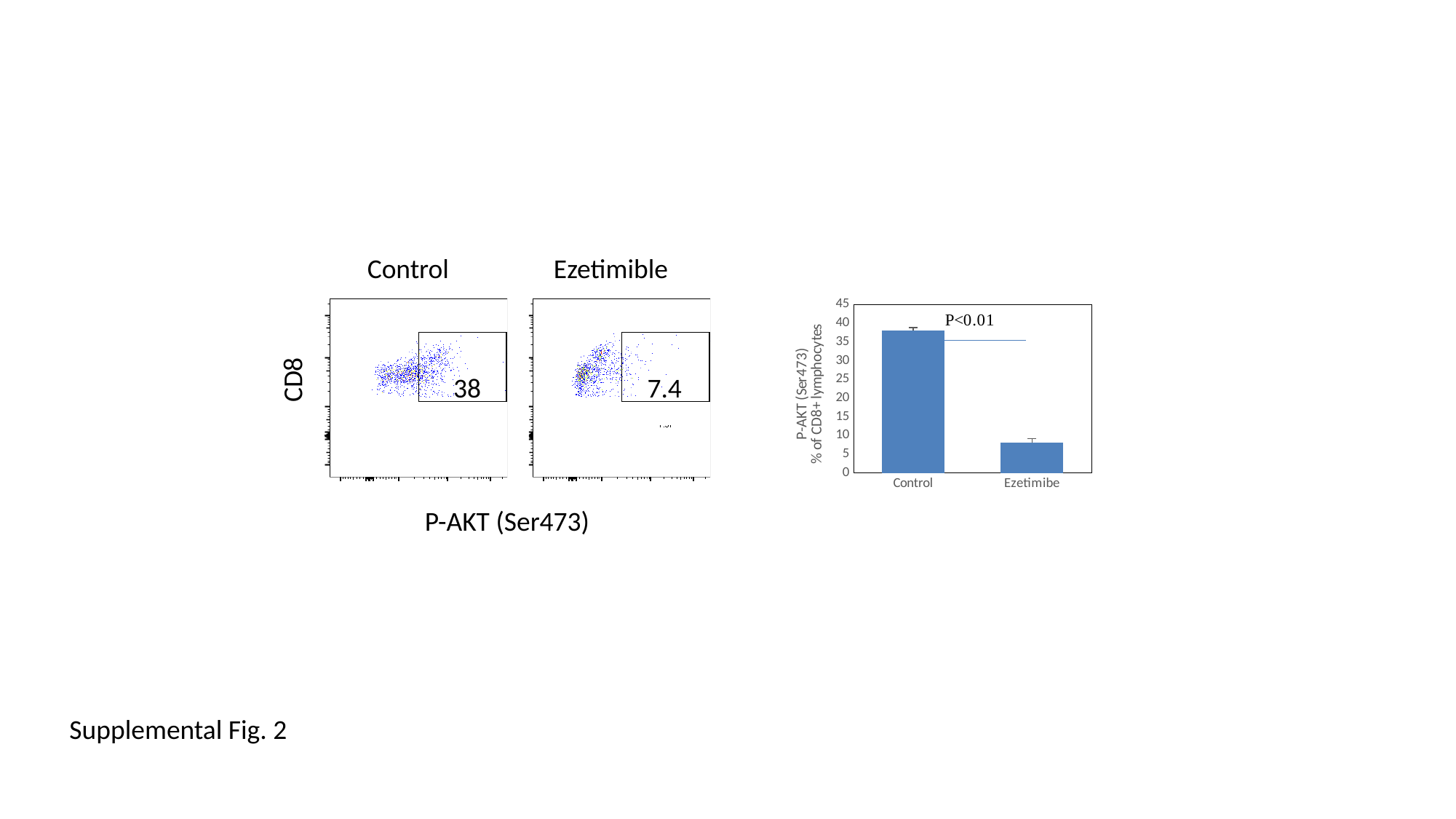

Control Ezetimible
#### Chart
| Category | |
|---|---|
| Control | 38.13333333333333 |
| Ezetimibe | 8.133333333333335 |CD8
38
7.4
P-AKT (Ser473)
Supplemental Fig. 2

### Slide 3
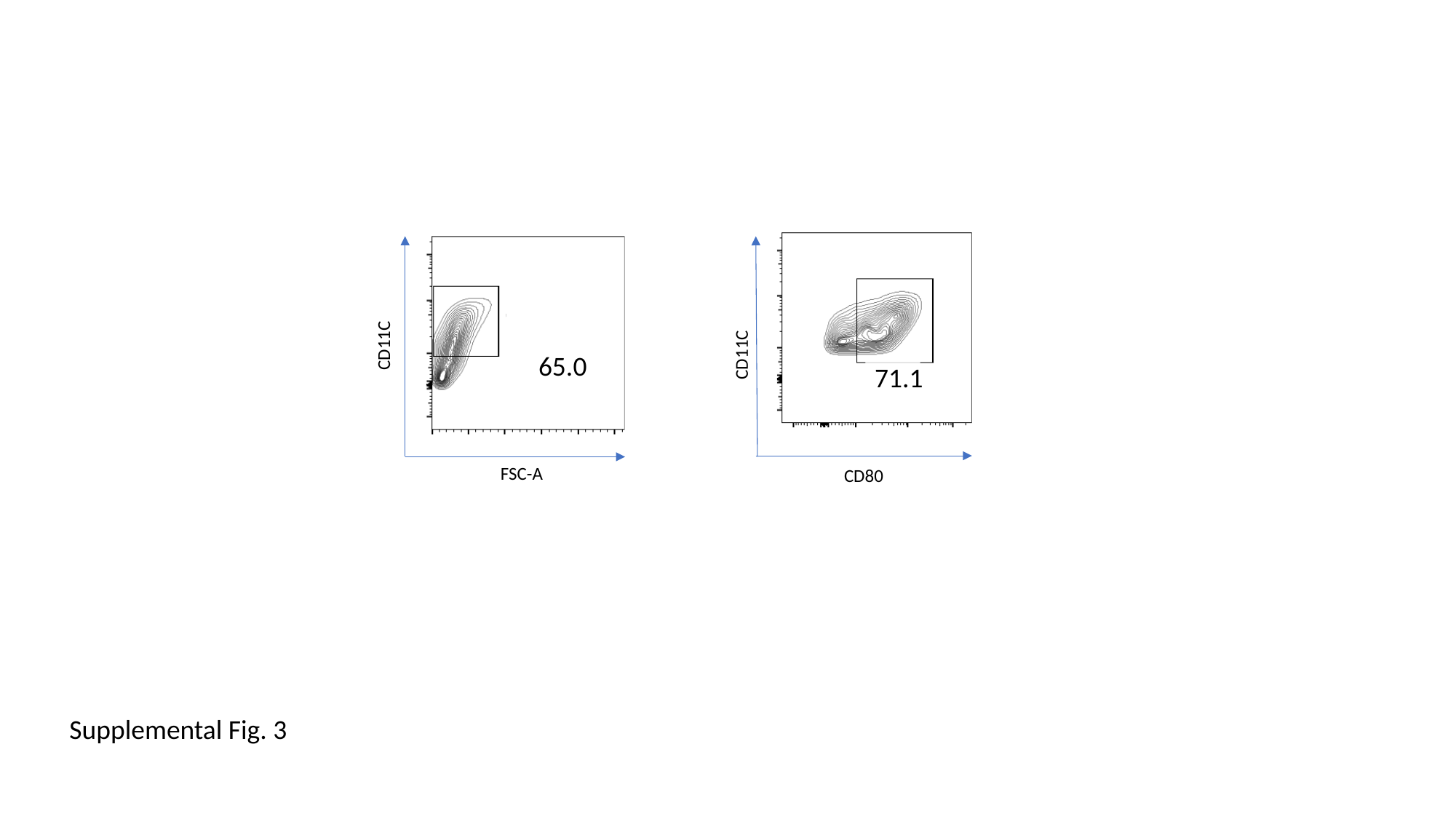

CD11C
CD11C
CD11C
65.0
71.1
6.28
FSC-A
CD80
CD86
Supplemental Fig. 3

### Slide 4
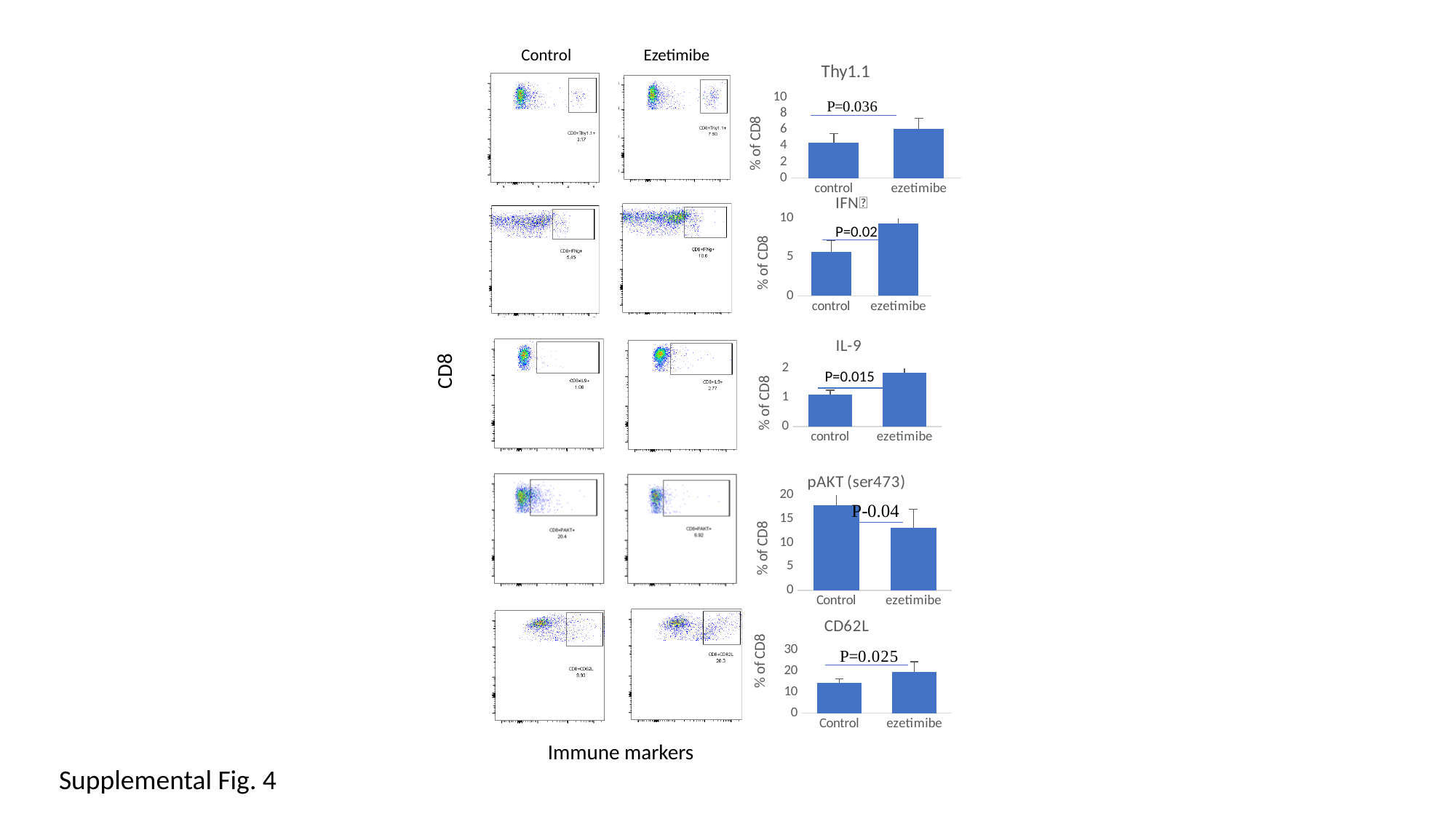

Control Ezetimibe
#### Chart: Thy1.1
| Category | |
|---|---|
| control | 4.3999999999999995 |
| ezetimibe | 6.1000000000000005 |
#### Chart: IFN
| Category | |
|---|---|
| control | 5.650000000000001 |
| ezetimibe | 9.366666666666665 |
P=0.02
#### Chart: IL-9
| Category | |
|---|---|
| control | 1.0899999999999999 |
| ezetimibe | 1.8500000000000003 |
CD8
P=0.015
#### Chart: pAKT (ser473)
| Category | |
|---|---|
| Control | 17.833333333333332 |
| ezetimibe | 13.15 |
#### Chart: CD62L
| Category | |
|---|---|
| Control | 14.200000000000003 |
| ezetimibe | 19.666666666666668 |
Immune markers
Supplemental Fig. 4

### Slide 5
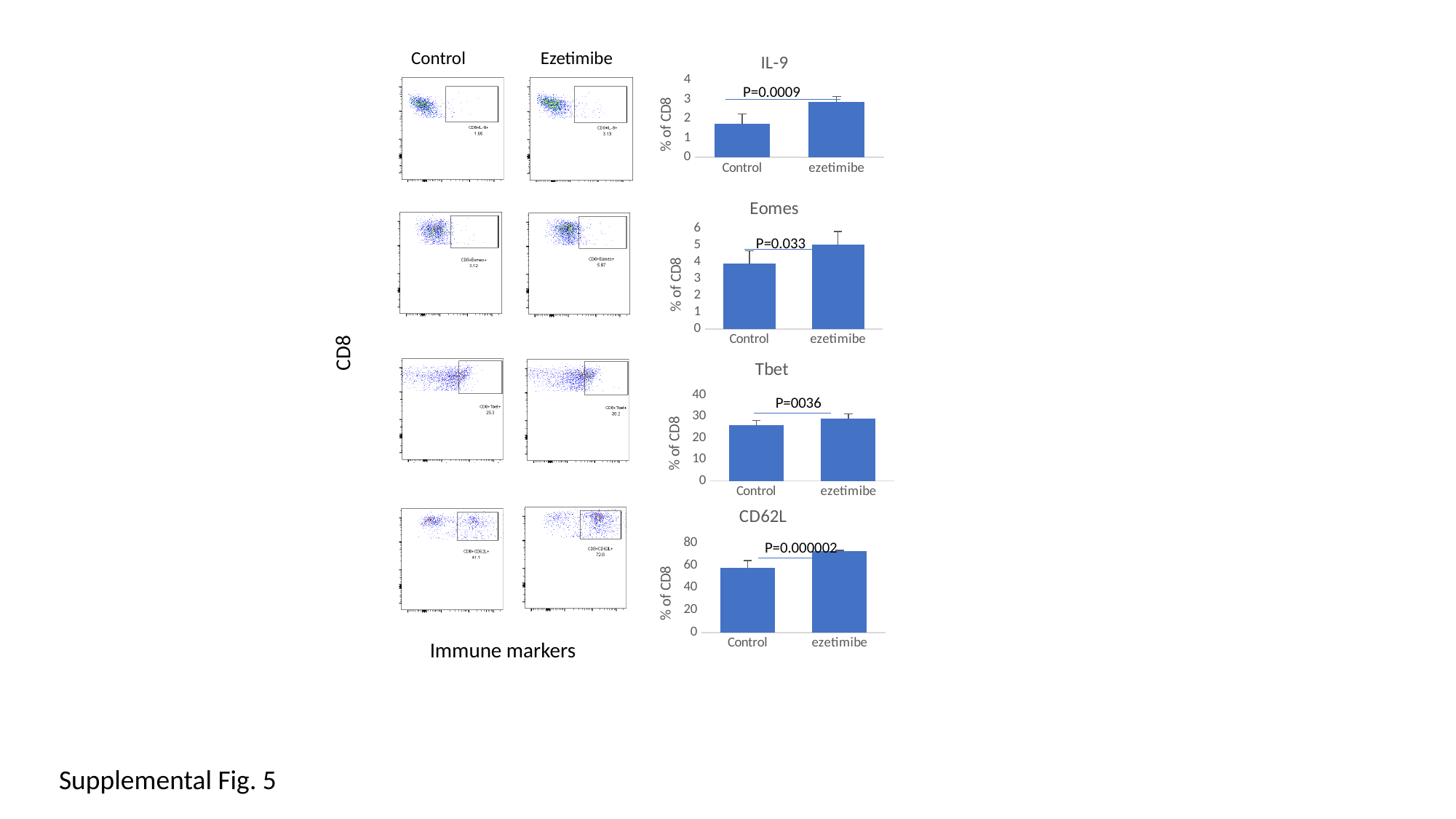

#### Chart: IL-9
| Category | |
|---|---|
| Control | 1.7333333333333334 |
| ezetimibe | 2.866666666666667 |Control Ezetimibe
P=0.0009
#### Chart: Eomes
| Category | |
|---|---|
| Control | 3.9166666666666674 |
| ezetimibe | 5.033333333333332 |
P=0.033
CD8
#### Chart: Tbet
| Category | |
|---|---|
| Control | 25.783333333333335 |
| ezetimibe | 28.900000000000002 |
P=0036
#### Chart: CD62L
| Category | |
|---|---|
| Control | 57.675 |
| ezetimibe | 72.28750000000001 |
P=0.000002
Immune markers
Supplemental Fig. 5

### Slide 6
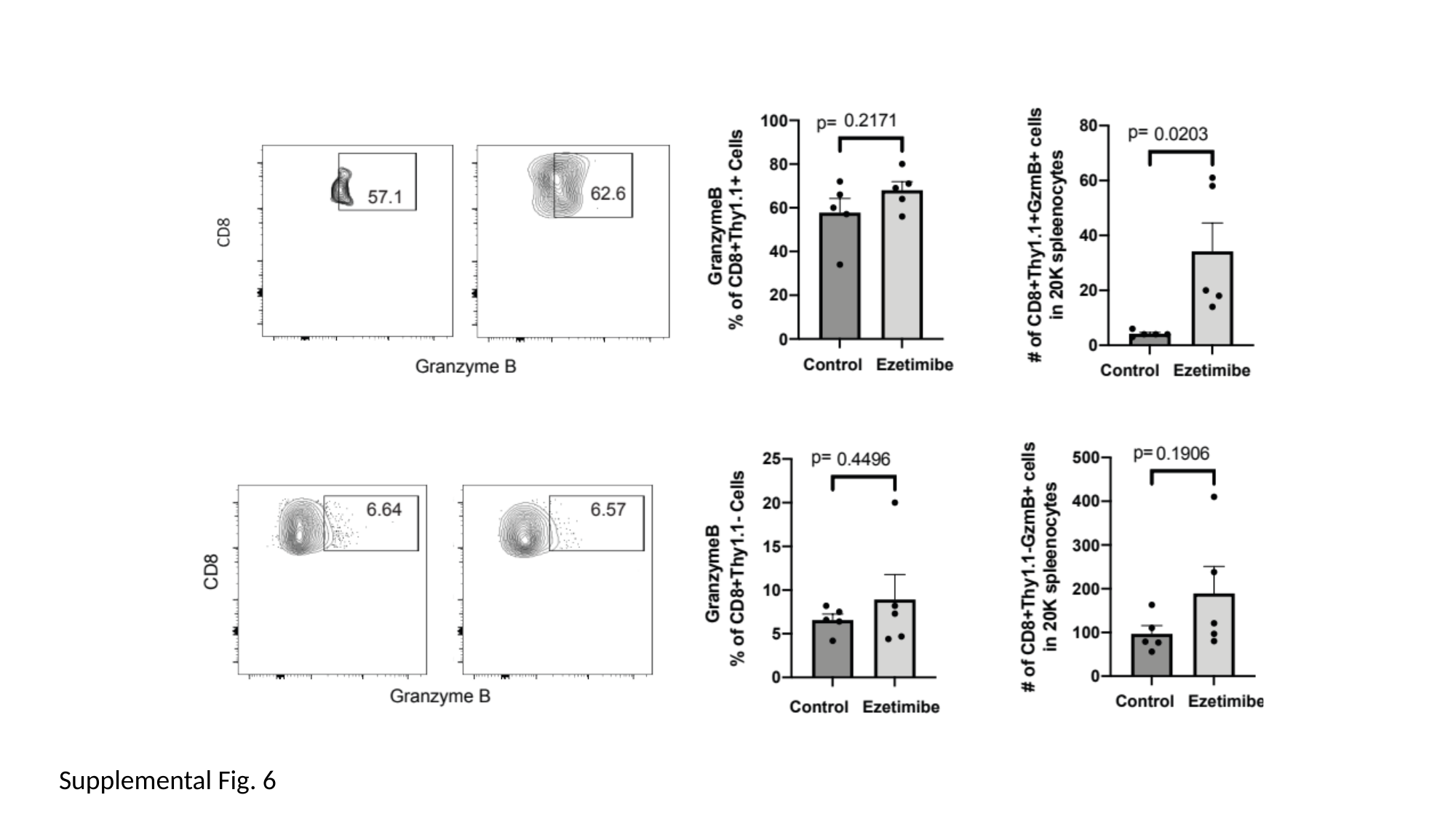

Supplemental Fig. 6

### Slide 7
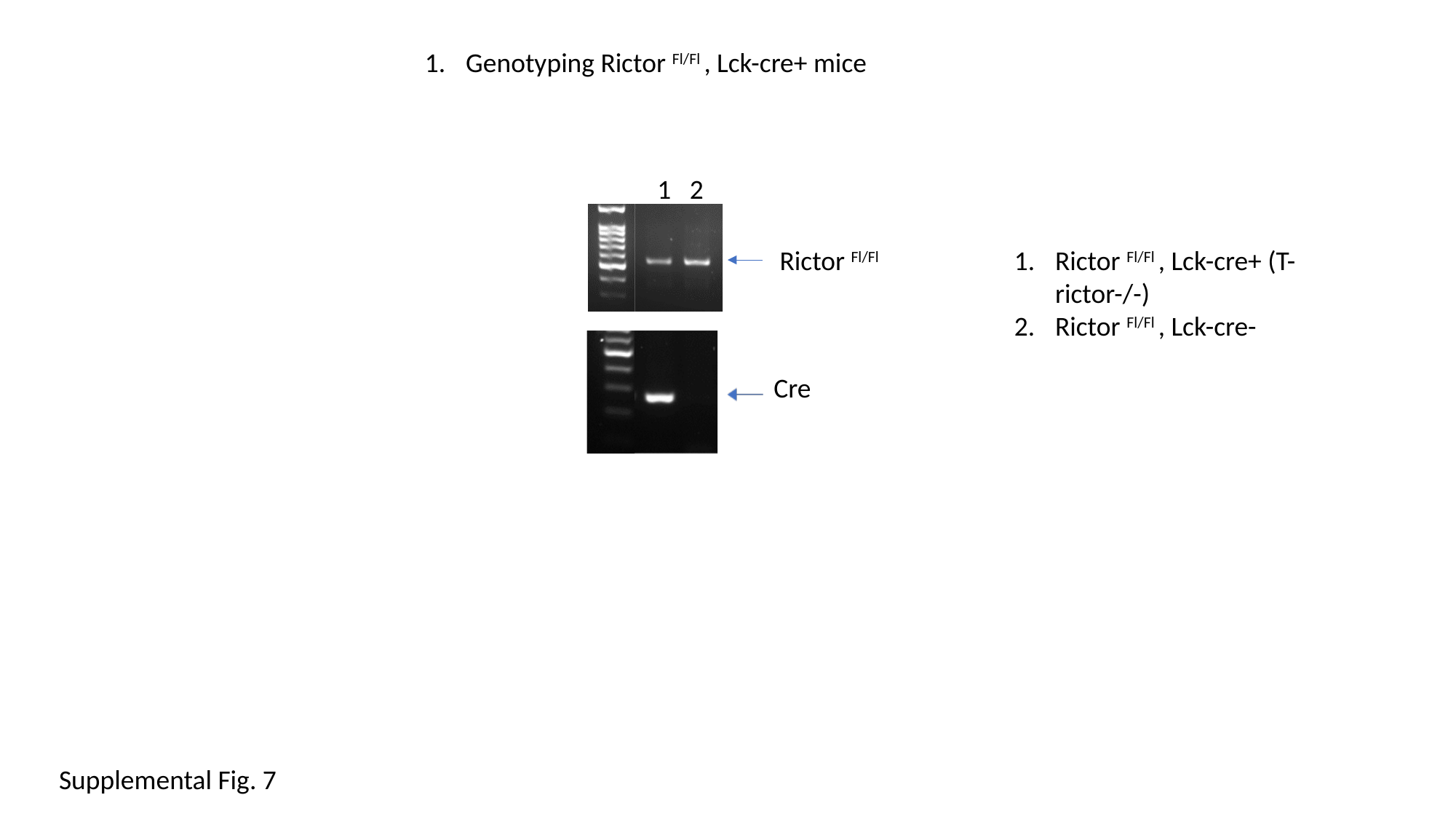

Genotyping Rictor ^Fl/Fl^ , Lck-cre+ mice
1 2
Rictor ^Fl/Fl^
Rictor ^Fl/Fl^ , Lck-cre+ (T-rictor-/-)
Rictor ^Fl/Fl^ , Lck-cre-
Cre
Supplemental Fig. 7

### Slide 8
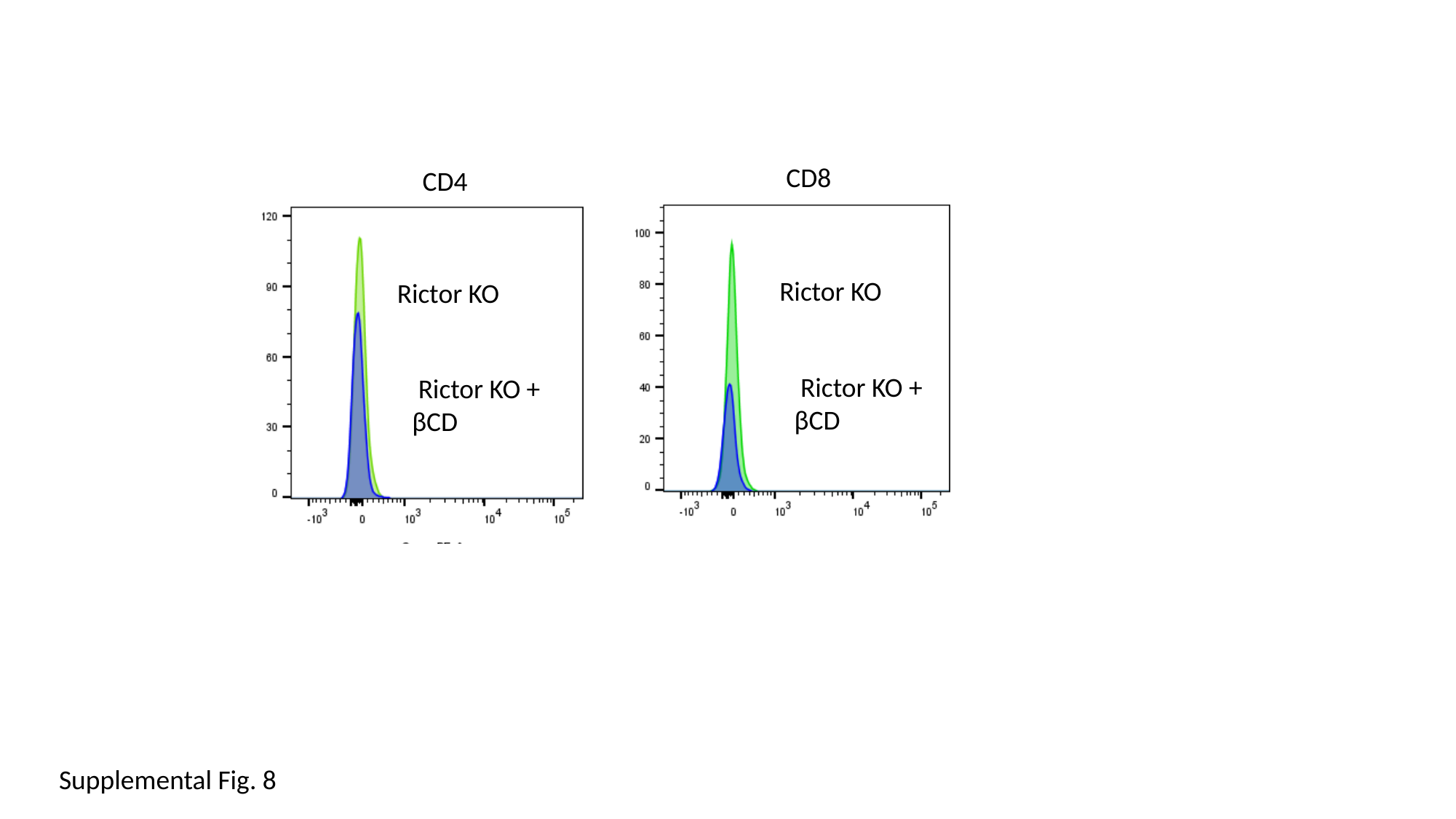

CD8
CD4
 Rictor KO
 Rictor KO
 Rictor KO +
βCD
 Rictor KO +
βCD
Supplemental Fig. 8

### Slide 9
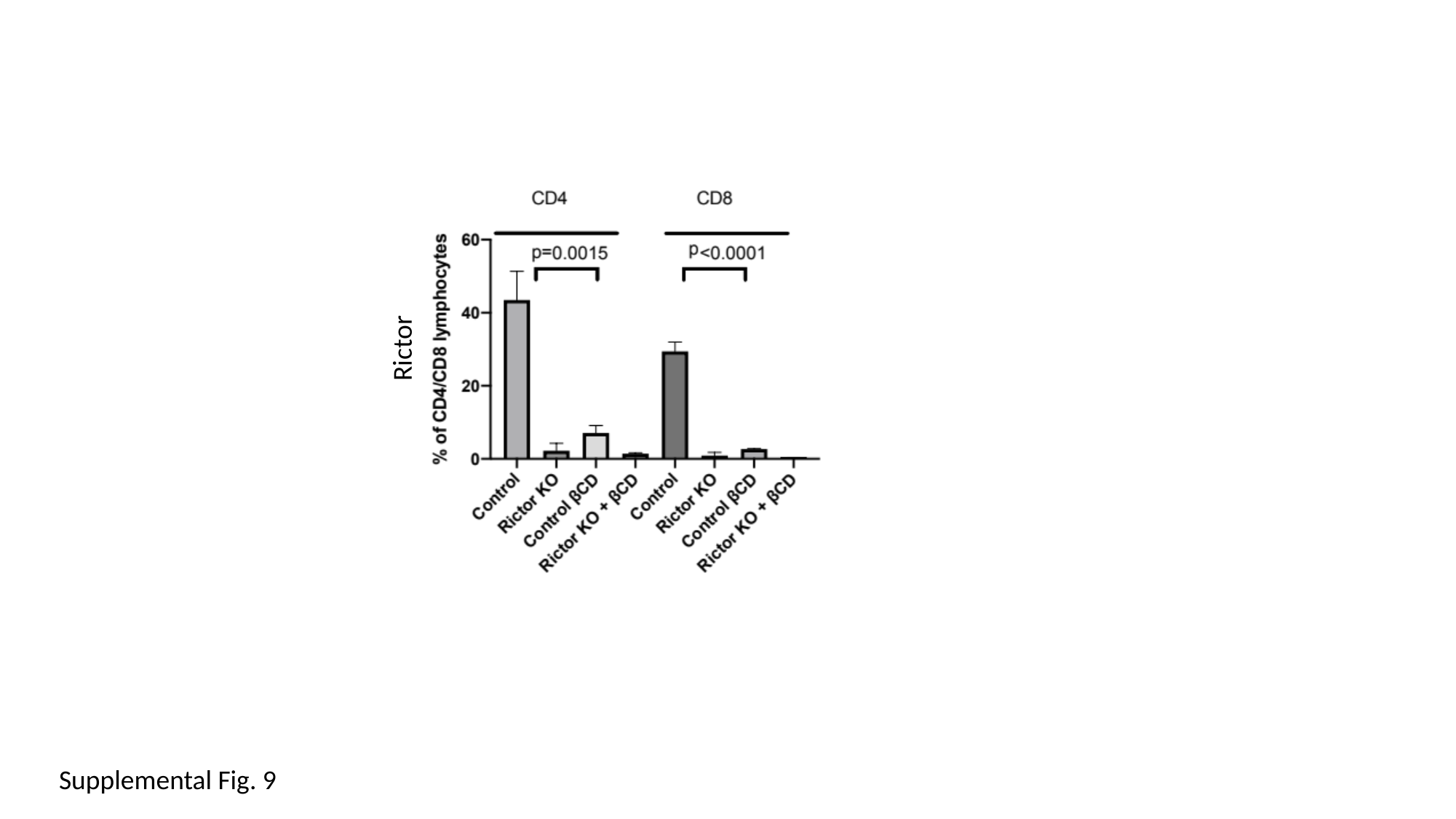

Rictor
Supplemental Fig. 9

### Slide 10
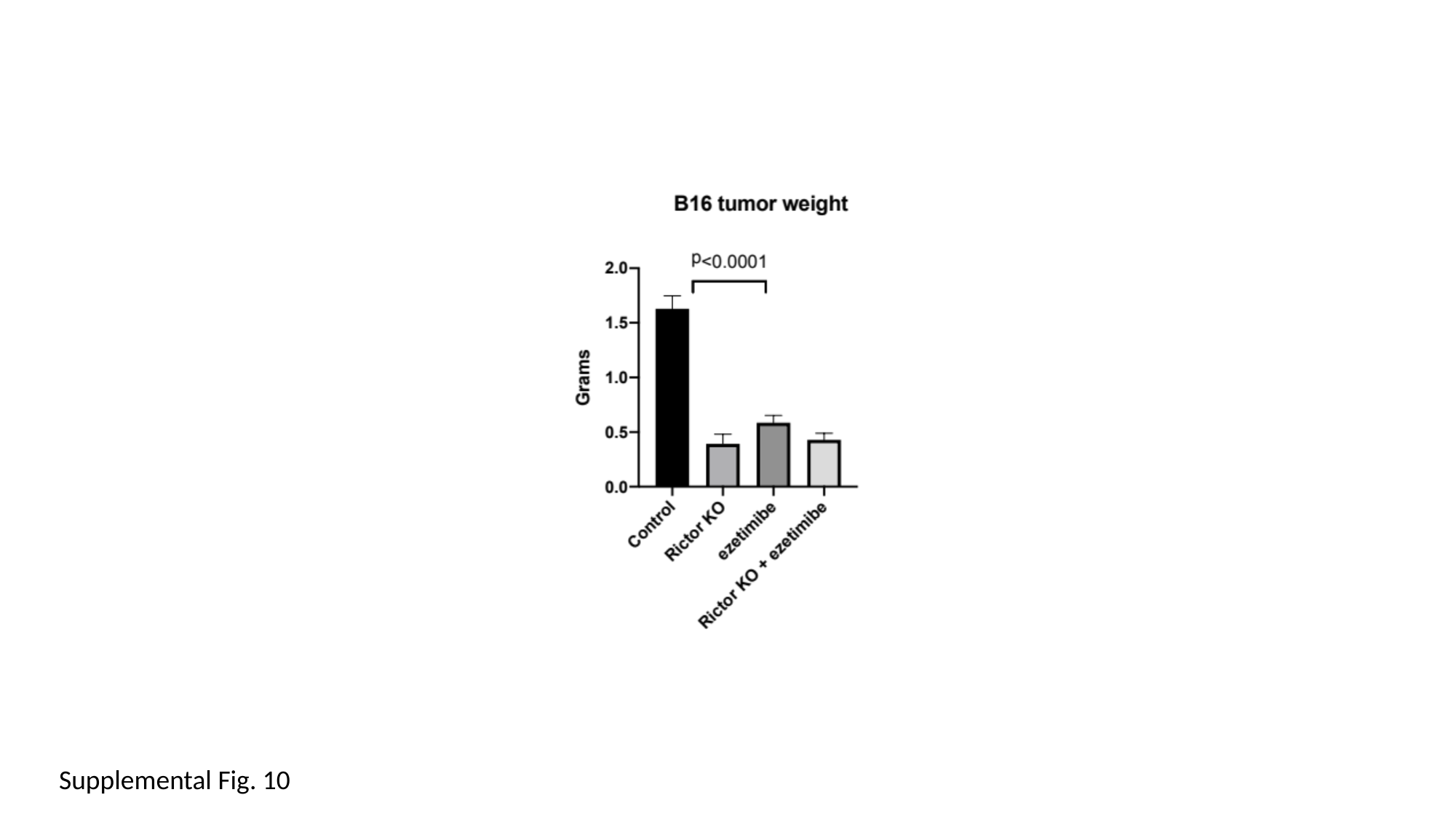

Supplemental Fig. 10

### Slide 11
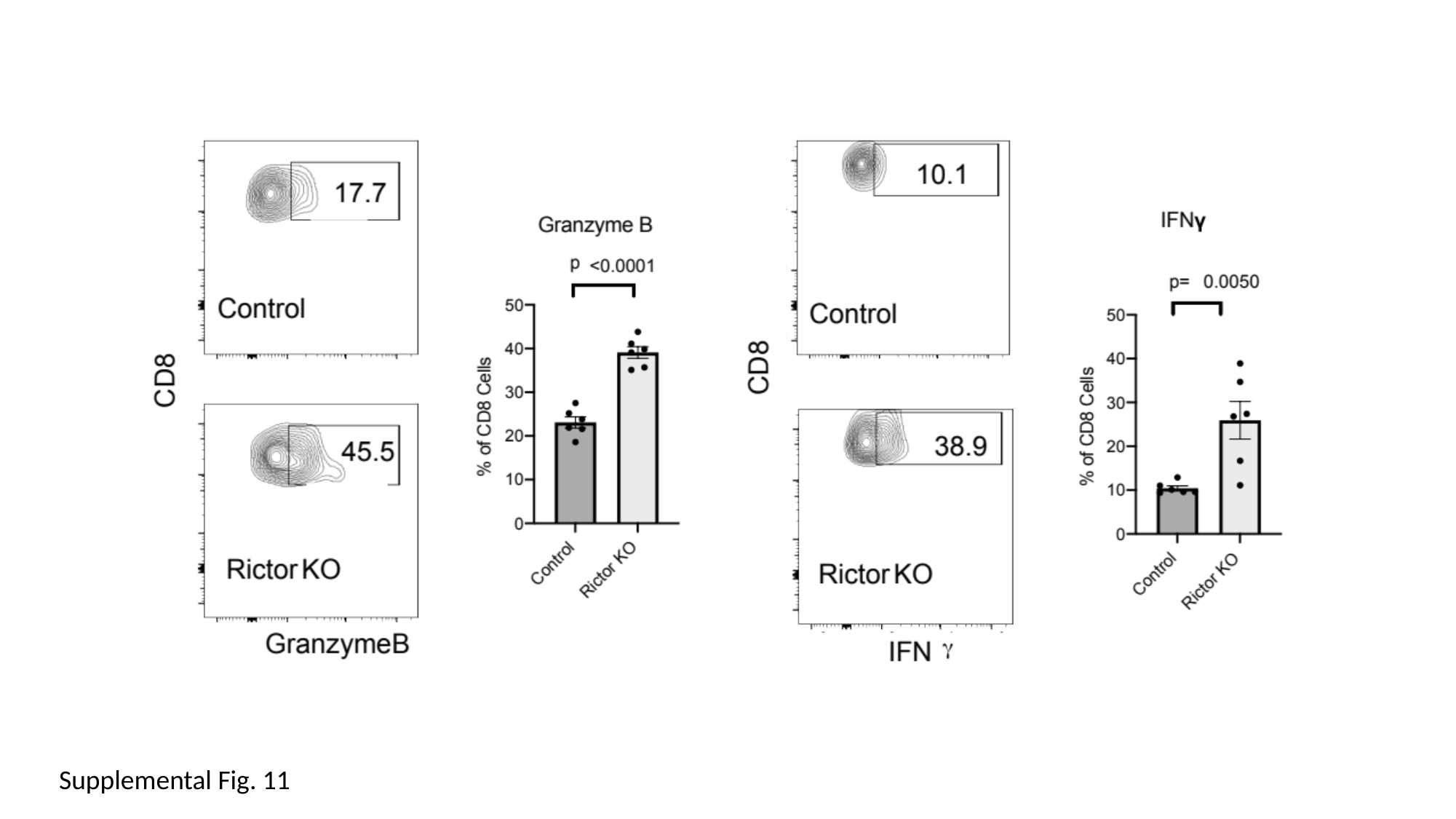

Supplemental Fig. 11

### Slide 12
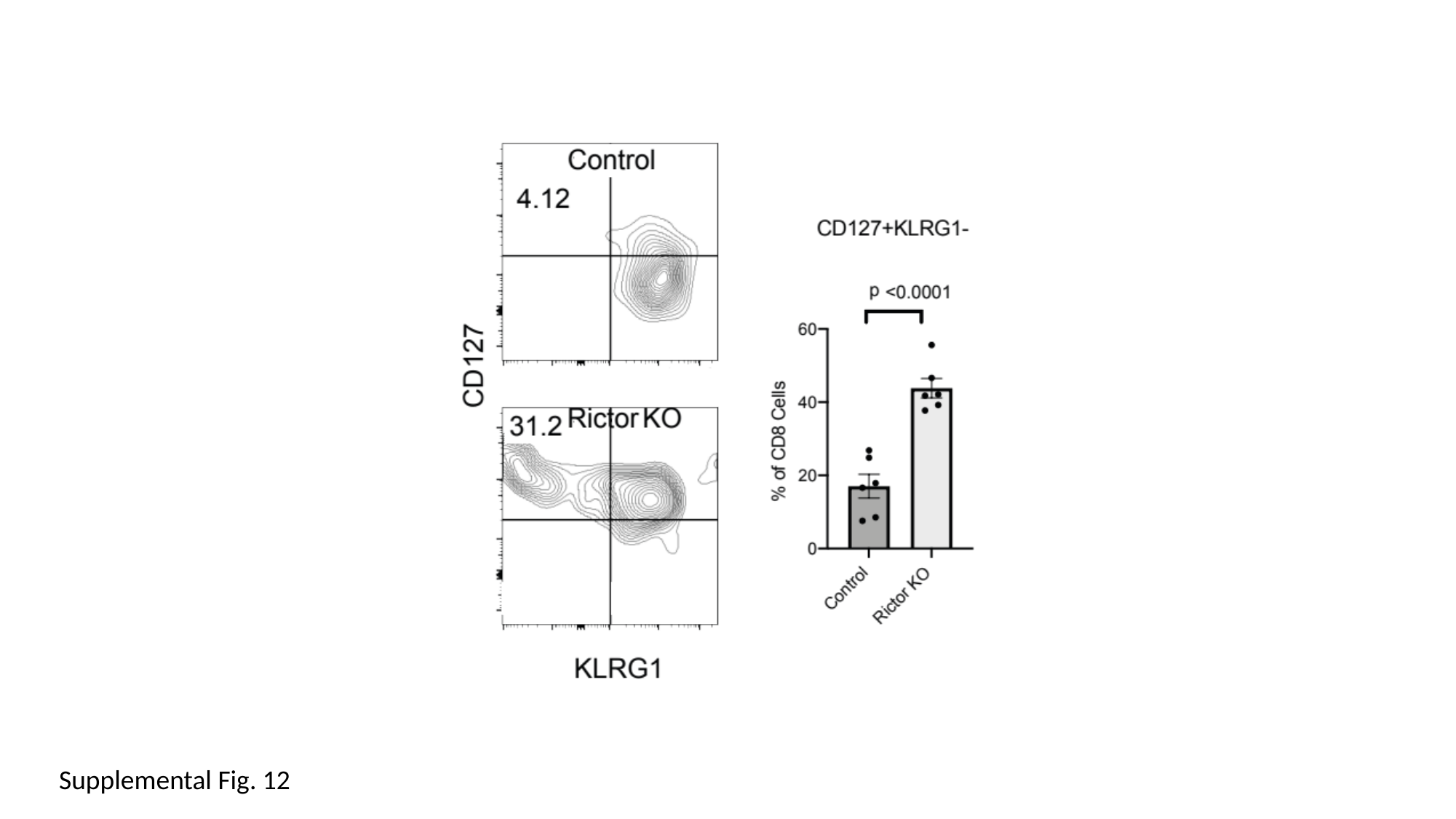

Supplemental Fig. 12
